# The tumour microenvironment influences long-term tamoxifen benefit in postmenopausal ER+/HER2- breast cancer patients

**DOI:** 10.64898/2026.03.24.26349151

**Authors:** Paula Camargo-Romera, Miguel Castresana-Aguirre, O. Danielsson, Huma Dar, Arne Östman, Kamila Czene, L. S. Lindström, Nicholas P. Tobin

## Abstract

**Background:** The tumour microenvironment (TME) influences breast cancer progression and treatment response. We investigated whether TME composition predicts tamoxifen benefit in postmenopausal women with oestrogen receptor-positive, HER2-negative (ER+HER2-) breast cancer.

**Methods:** This study included 513 patients from the Stockholm Tamoxifen (STO-3) trial, which randomised postmenopausal, lymph node–negative women to tamoxifen or no endocrine therapy. Bulk tumour transcriptomes were deconvoluted with the ConsensusTME algorithm to estimate the relative abundance of 18 immune and stromal cell types. A summary score of combined immune cells was created on a per patient basis and evaluated alongside fibroblast and endothelial stromal compartments. Patients were categorised into immune and stromal tertiles on the basis of these scores. Associations between TME composition and tumour characteristics were evaluated using Spearman correlations and Fisher’s exact test. Tamoxifen benefit was analysed by univariable Kaplan-Meier (log-rank) and multivariable Cox proportional hazards adjusting for age, tumour size, grade, progesterone receptor, Ki-67, and radiotherapy. Differential expression was assessed with limma and pathway enrichment with fgsea using Hallmark gene sets from MSigDB.

**Results:** Low immune abundance was significantly associated with higher ER expression (Fisher’s exact test p < 0.001). Among tamoxifen-treated patients, those with low immune scores showed improved distant recurrence-free interval (DRFI) relative to untreated patients (log-rank p < 0.001). Similarly, intermediate endothelial (p < 0.001) and low/intermediate fibroblast abundances (p = 0.042, p = 0.009) were associated with favourable DRFI. In multivariable models, low immune (aHR = 0.17, 95% CI 0.08–0.40), intermediate endothelial (aHR = 0.21, 95% CI 0.09–0.51), and low/intermediate fibroblast tertiles (aHR = 0.50, 95% CI 0.27–0.93; aHR = 0.36, 95% CI 0.17–0.77) retained significance. Transcriptomic analysis revealed enrichment of oestrogen-response, MYC-target, and oxidative-phosphorylation pathways in low-immune and low-fibroblast tumours, while interferon-γ response and allograft rejection pathways were downregulated.

**Conclusions:** TME composition modulates tamoxifen benefit in postmenopausal ER+HER2-breast cancer. Low immune, intermediate endothelial, and low/intermediate fibroblast abundances are associated with improved benefit from tamoxifen, suggesting that both immune and stromal compartments influence endocrine treatment efficacy.

## INTRODUCTION

Breast cancer is one of the most common types of female cancers and is a principal cause of mortality in women globally(1,2). Understanding the biology of breast cancer poses a challenge owing to its heterogeneous nature, where individual tumours exhibit diverse characteristics and behaviours(3). This complexity is further compounded by the influence of the tumour microenvironment (TME), making it challenging to unravel disease mechanisms and develop effective treatment strategies(3).

The TME is comprised of tumour, immune and stromal cells along with the extracellular matrix (a non-cellular component)(4). It plays a crucial role in breast cancer proliferation, immune system suppression, response to treatment(5) and disease progression(6). Importantly, the cellular composition and abundance of TME cells have been shown to vary depending on tumour oestrogen receptor status(7) and this variability has been shown to influence the survival of breast cancer patients. In ER-positive tumours with up to 10 years of median follow-up, patients with a higher abundance of M0 macrophages show poorer survival than those with lower abundances(8). Conversely, in ER-negative tumours, the presence of CD8+ and activated memory T cells have been associated better survival outcomes(8–11). How these subtypes influence the long term (>25 year) survival of breast cancer patients is currently unclear.

These studies highlight the importance of examining the TME separately on the basis of ER status. To date however, studies relating TME cell abundance to treatment response in randomised clinical trial samples from ER-positive tumours are lacking. Here, we perform a comprehensive analysis of how tumour microenvironment (TME) composition influences the benefit derived from tamoxifen treatment in ER-positive, HER2-negative postmenopausal breast cancer patients within the randomised STO-3 trial. This trial uniquely includes a no-endocrine-therapy control arm enabling direct comparison of treatment benefit. We apply a deconvolution technique to determine the relative abundance of eighteen TME cell types and perform uni- and multi-variable survival analyses to determine their impact on tamoxifen-treated patient survival relative to non-tamoxifen treated patients.

## METHODS

### The Stockholm Tamoxifen Clinical Trial

The Stockholm Breast Cancer Study Group conducted randomised trials from 1976 and onwards(12,13) . The Stockholm Tamoxifen Trial 3 (STO-3) enrolled 1780 postmenopausal patients with lymph node-negative breast cancer who were randomised to two years of adjuvant tamoxifen (40 mg daily) or no adjuvant treatment(12,13). In 1983, patients who re-consented and were disease-free after 2 years of tamoxifen treatment were randomised to 3 more years of tamoxifen or no further treatment(12,13). A subset of 808 patients with formalin-fixed paraffin-embedded (FFPE) material was available for molecular analyses, and this patient subset is well-balanced relative to the original STO-3 trial regarding tumour characteristics(13,14). Samples from eighty-one of these patients were excluded due to insufficient invasive tumour cells, leaving 727 samples in total(14) (Figure 1). Comprehensive patient and clinicopathological annotations were present for all participants(13). Follow-up until December 31, 2016, was obtained from Swedish national and regional registries of high validity and essentially complete coverage(15–17) . Detailed patient and clinical information was available for all patients in the trials. The STO-trials were approved by the Karolinska Institutet Regional Ethics Committee with the Stockholm Regional Cancer Center as the trial center. Informed consent was obtained before randomization. The trials were approved and initiated before the practice of trial registration in Sweden(15–17).

**Figure 1.**
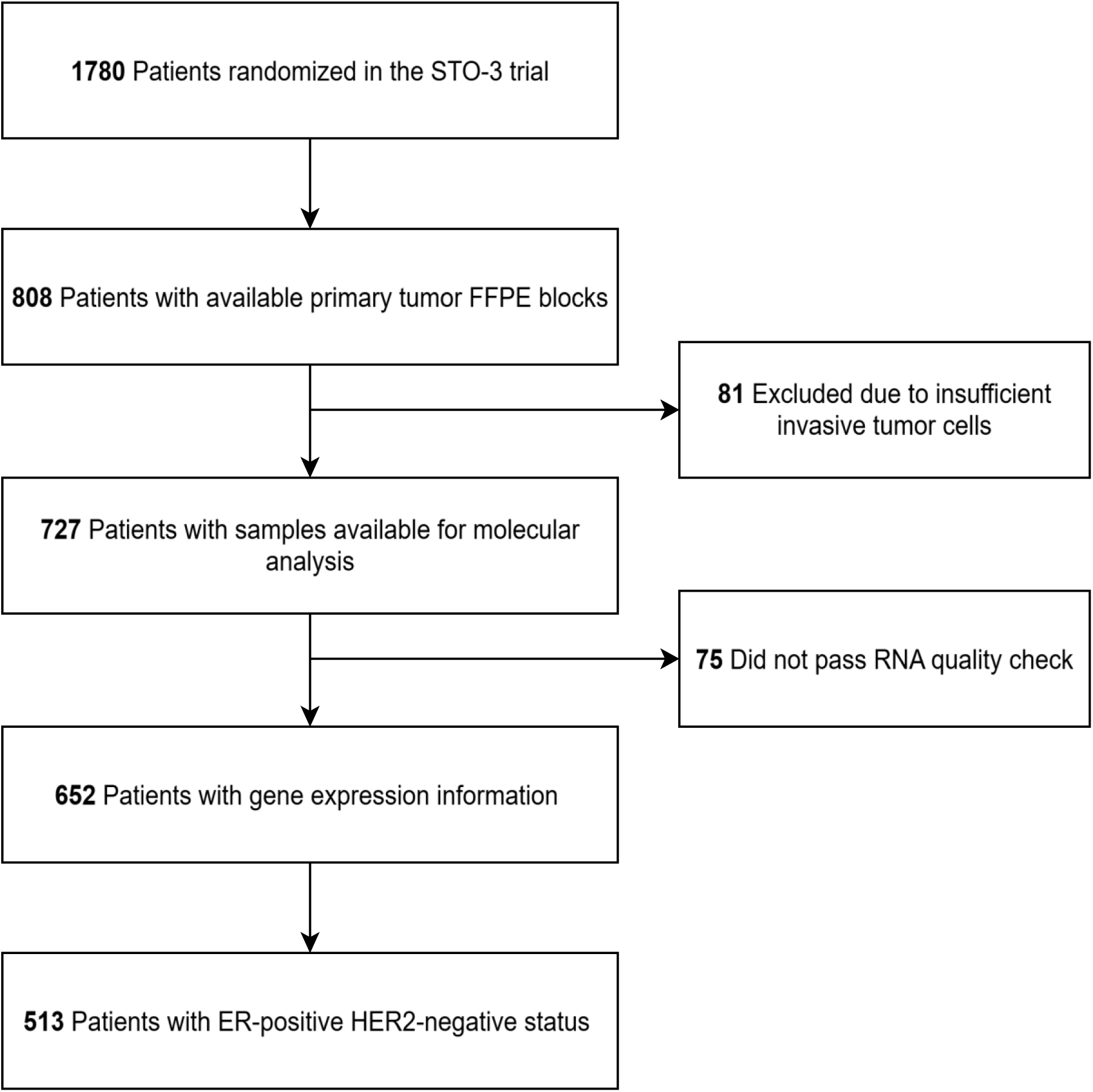
CONSORT diagram of the Stockholm tamoxifen randomised trial (STO-3 trial). ER, oestrogen receptor. HER2, human epidermal growth factor receptor 2.

### Immunochemistry

Immunohistochemical analysis for ER, PR, HER2, and Ki-67 was performed in 2014 (STO-3).(13) The percentage of cancer cells positive for ER, PR, HER2, and Ki-67 was scored by experienced breast cancer pathologists. ER-positive and PR-positive status were defined by a threshold of 10% or greater according to Swedish National guidelines (18), and Ki-67 was measured in the whole tumour section and categorized as low (<15%) and intermediate/high (≥15%). HER2 positivity was defined as intensity 3+ by IHC.

### Tumour Size and Tumour Grade

Tumour diameter was measured according to clinical guidelines and classified into three groups: T1a/b (≤ 10 mm), T1c (11-20 mm), and T2-T3 (> 20 mm). Tumour grade was assessed according to the Nottingham Histological Score system (Elston grade)(19).

### Agilent microarray gene expression profiling

Agilent microarray profiling was performed on FFPE breast cancer tumour tissue in 2014(13) using custom-designed arrays from Agilent Technologies (Santa Clara, CA). Each array contained ∼32.1K probes representing ∼21.5K unique genes. Of the 727 breast cancer tumours examined, 652 successfully passed the RNA quality check according to the diagnostic quality model and out of these, 513 tumours were ER-positive/HER2-negative (Figure 1).

### Deconvolution of Tumour Microenvironment and Immune Score Construction

The deconvolution algorithm Consensus^TME^ (20) was employed to discern the composition of TME cell types from bulk microarray data and was chosen owing to its cancer specificity and integration of multiple methodologies. The algorithm outputs normalized enrichment score (NES) scores on a per tumour basis that reflects the relative abundance of 18 TME cell types. These include Regulatory T cells, γδ T cells, B cells, CD4+ T cells, CD8+ T cells, Cytotoxic cells, Dendritic cells, Eosinophils, Macrophages, M1 macrophages, M2 macrophages, Mast cells, Monocytes, Neutrophils, NK cells, Plasma cells, Fibroblasts, and Endothelial cells.

As ConsensusTME estimates are comparable across patients but not between cell types, we first applied z-score normalisation to each cell type across all patients. The resulting standardised scores were then summed across all immune cell types (e.g. all cell types noted above with the exception of fibroblasts, and endothelial cells) within each tumour sample to construct an aggregated immune score. This continuous variable was subsequently split into tertiles of low, intermediate and high relative immune abundance and used for survival and differential gene expression analyses. The same tertile split was applied to the continuous NES scores for fibroblast and endothelial cell abundances. Finally, a semi-supervised hierarchical clustering was performed using the average agglomeration method (UPGMA) to indicate similarity in immune abundance between samples clustering columns on the basis of immune score tertiles and rows on the basis of Pearson correlation.

### Differential expression and gene set enrichment analysis

To examine genes and pathways associated with distinct tumour microenvironment cell type compositions, we performed differential gene expression and pathway enrichment analyses based on the immune, endothelial, and fibroblast tertiles. For the immune and fibroblast scores, samples exhibiting low vs high relative cell-type abundance were compared to capture genes differentially expressed between these extremes. For the endothelial score, the intermediate group was compared to low and high separately, in line with results from survival analyses (see Results). Differential expression analysis was performed using the limma package (version 3.62.2) in R, employing empirical Bayes moderation to estimate log2 fold changes and associated statistical significance. Genes were subsequently ranked by their moderated t-statistics. To ensure that differentially expressed genes did not substantially overlap with genes used to define Consensus^TME^ cell-type signatures, we calculated the Jaccard similarity index between these sets. Pathway enrichment analysis was carried out using the fgsea package (version 1.32.4), with curated gene sets sourced from the MSigDB database via the msigdbr package (version 10.0.1). Specifically, the Hallmark gene sets (collection ’H’) were utilized to identify key biological pathways enriched among the differentially expressed genes. Enrichment significance was assessed using the Benjamini–Hochberg method to control the false discovery rate (FDR).

### Statistical analysis

To test for correlations between NES scores and TME abundances, we performed a Spearman correlation analysis, clustering based on the average agglomeration method (UPGMA), with p < 0.05 and a two-sided alternative hypothesis. Fisher’s exact test was used to assess differences in patient and tumour characteristics between Immune, Endothelial and Fibroblast tertiles. Differences in distance recurrence between tamoxifen treated vs. non-treated were examined within immune, endothelial and fibroblast tertiles by univariable Kaplan-Meier (log-rank test) and multivariable Cox proportional hazard regression (Wald test) analyses. These analyses were designed to estimate tamoxifen benefit within each TME tertile, rather than overall prognostic effects, by directly comparing treated versus untreated patients. The clinical endpoint of interest for all analyses was distant recurrence-free interval (DRFI), with distant disease, i.e. distant metastatic disease, as the event(21). Follow-up started at the date of primary breast cancer diagnosis and ended at the date of distant recurrence, date of death, or end of study follow-up (December 31, 2016). Multivariable analyses were adjusted for patient and tumour characteristics, including age, tumour size, tumour grade, progesterone receptor status, Ki-67 status, and the administration of radiotherapy. Interaction between the different scores and tamoxifen was tested by including a product term in the Cox proportional hazard regression model. All data preparation and analyses were performed using R version 4.4.2. Kaplan-Meier and Cox proportional hazard analyses were assessed using the R survival package, the R survminer package was used for survival curves, and the pheatmap package was used for hierarchical clustering analyses. All statistical tests were two-sided; statistical significance was assessed at α = 0.05,

## RESULTS

### Characterization of the Tumour Microenvironment

A total of 513 ER+ HER- postmenopausal women from the STO-3 trial with associated microarray-based transcriptomic data were included in this study (Figure 1). No significant differences in patient and tumour characteristics were observed between those who received tamoxifen treatment and those who did not (Table 1).

**Table 1.** Patient and Tumour Characteristics for 513 Oestrogen Receptor-Positive and HER2 Receptor-Negative Patients in STO-3.

|  |  | Tamoxifen Untreated (n = 239) | Tamoxifen Treated (n = 274) | p<sup>a</sup> |
| --- | --- | --- | --- | --- |
| Age, N (%) | 45-54 | 25 (10.5) | 21 (7.7) | 0.552 |
|  | 55-64 | 118 (49.4) | 139 (50.7) |  |
|  | 65-74 | 96 (40.1) | 114 (41.6) |  |
| Tumour size, N (%) | <=20 mm | 191 (79.9) | 226 (82.5) | 0.746 |
|  | >20 mm | 45 (18.8) | 45 (16.4) |  |
|  | Unknown | 3 (1.3) | 3 (1.1) |  |
| Tumour grade, N (%) | 1 | 59 (24.7) | 56 (20.4) | 0.609 |
|  | 2 | 146 (61.1) | 180 (65.7) |  |
|  | 3 | 31 (13.0) | 33 (12.1) |  |
|  | Unknown | 3 (1.2) | 5 (1.8) |  |
| PR status, N (%) | Positive | 170 (71.1) | 191 (69.7) | 0.928 |
|  | Negative | 66 (27.6) | 80 (29.2) |  |
|  | Unknown | 3 (1.3) | 3 (1.1) |  |
| Ki-67 status, N (%) | Positive | 55 (23.1) | 50 (18.3) | 0.209 |
|  | Negative | 176 (73.6) | 208 (75.9) |  |
|  | Unknown | 8 (3.3) | 16 (5.8) |  |
| Radiotherapy, N (%) | Yes | 55 (23.0) | 63 (23.0) | 1.000 |
|  | No | 184 (77.0) | 211 (77.0) |  |

<sup>a</sup>p-value calculated by Fisher's exact test

The Consensus^TME^ deconvolution was applied to the bulk tumour microarray data and yielded normalized enrichment scale scores (NES) for 18 immune and stromal cell types of the tumour microenvironment on a per patient basis. In order to understand how these cell types relate to each other, we first performed a correlation analysis across all samples. We found that whilst in general immune cell types were significantly and intermediate to strongly correlated with each other (Spearman correlation > 0.5 for most comparisons, p < 0.05), no strong correlations were found between stromal cell types (endothelial and fibroblasts) and immune cell types (Figure 2A, Spearman correlation < 0.4 for all stromal vs. immune comparisons). Similarly, an unsupervised clustering of all samples showed immune cell types grouping separately from stromal cell types (Additional file 1, see dendrogram on the left-hand side, red arrows indicate stromal cell types). Based on these results, we chose to analyse immune cell types together and separately from their stromal counterparts.

**Figure 2.**
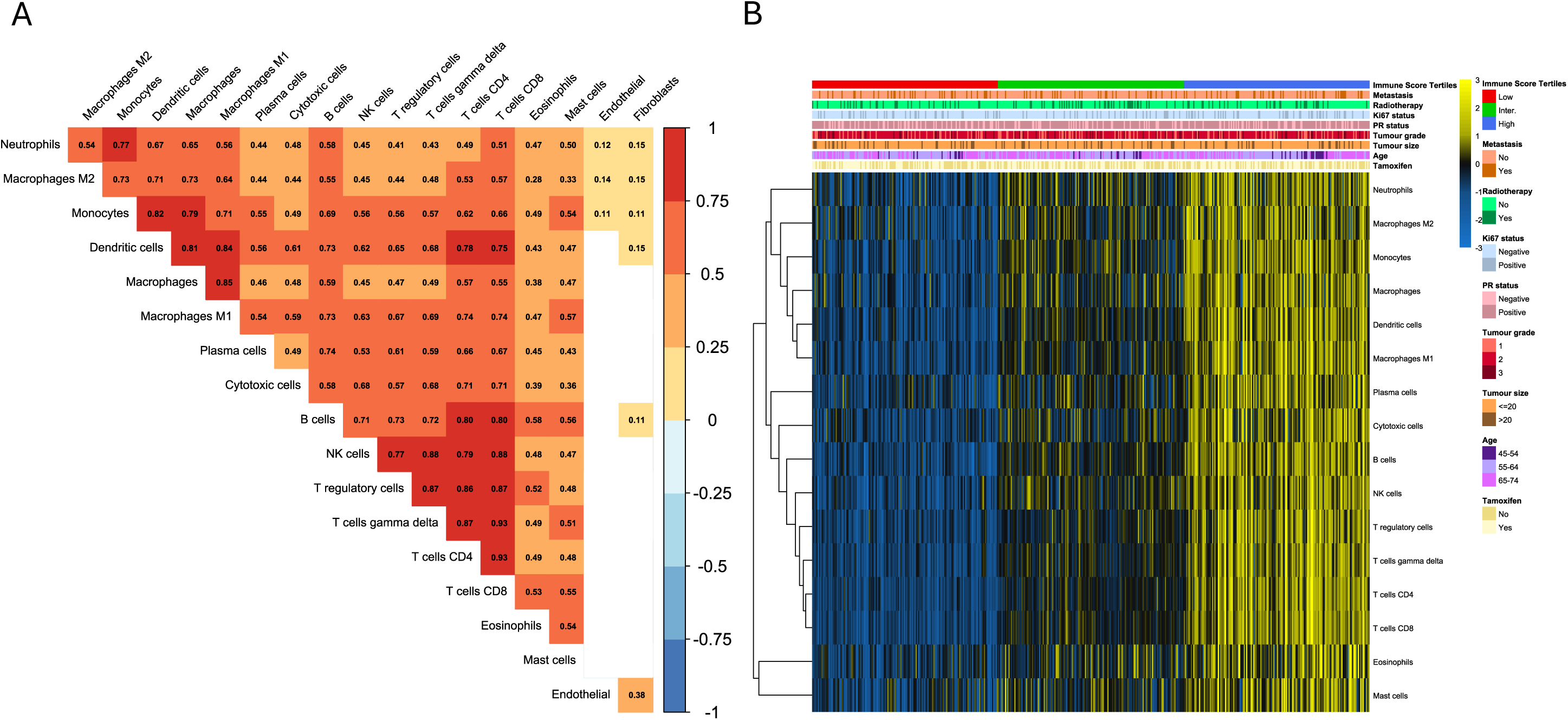
(A) Spearman correlation plot of 18 TME cell types, with coefficients ranging from -1 (negative) to 1 (positive); empty cells indicate non-significant correlations. (B) Semi-supervised heatmap of immune cell types across patient samples, ordered by immune score tertiles. The colour gradient (blue to yellow) reflects relative abundance. Top annotations show clinical and molecular features: metastasis, radiotherapy, Ki-67, PR status, grade, age, and tamoxifen treatment.

Given the high correlation between immune cell types, we next constructed an aggregated immune score by summing the values for all immune cell types and dividing the resulting continuous variable into tertiles of low, intermediate and high immune expression. A semi-supervised heatmap of immune cell types ordered by tertiles is shown in Figure 2B where again the similarities in immune cell expression within each tertile are readily apparent (Figure 2B, tertile proportions shown highest up on column legend). Tertiles were similarly constructed for endothelial and fibroblast continuous variables separately. When examining tumour characteristics in relation to these tertile groups, we found a significantly higher percentage of ER-positive cells in the immune score low tertile (Kruskal-Wallis test, p < 0.001, Table 2). Furthermore, statistically significant differences were noted between endothelial and fibroblast tertiles and tumour size, grade, Ki-67 and clinical risk (Table 2, Fisher’s exact test, p < 0.05). In general, low levels of both endothelial and fibroblast cell types were associated with more aggressive tumour characteristics (e.g. larger tumour size, grade 3 status, higher Ki-67 and higher clinical risk(22)).

**Table 2.**
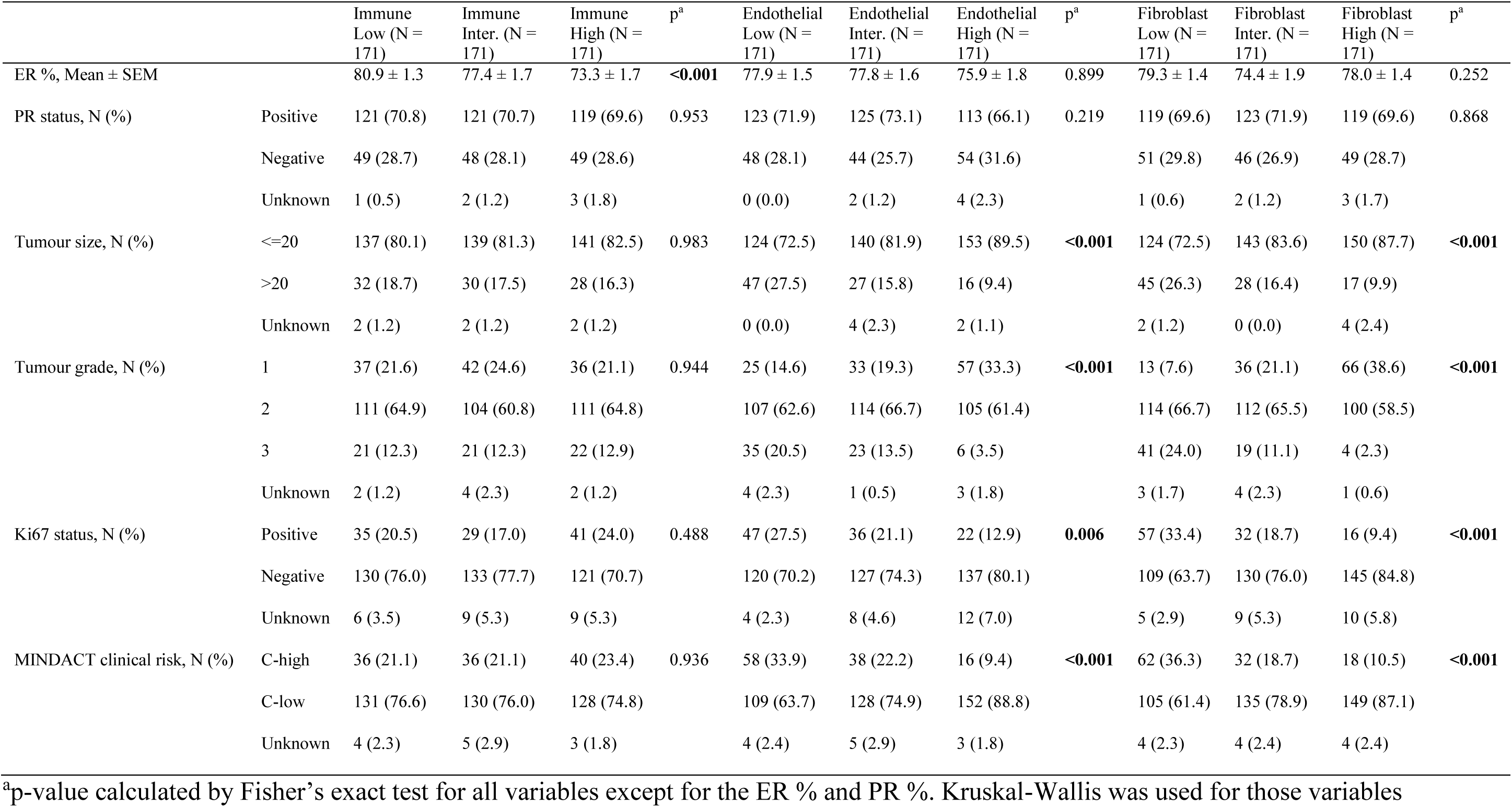
Tumour characteristics for STO-3 between the tertiles of Immune, Endothelial and Fibroblast scores.

|  |  | Immune<br>Low (N =<br>171) | Immune<br>Inter. (N =<br>171) | Immune<br>High (N =<br>171) | p<sup>a</sup> | Endothelial<br>Low (N =<br>171) | Endothelial<br>Inter. (N =<br>171) | Endothelial<br>High (N =<br>171) | p<sup>a</sup> | Fibroblast<br>Low (N =<br>171) | Fibroblast<br>Inter. (N =<br>171) | Fibroblast<br>High (N =<br>171) | p<sup>a</sup> |
| --- | --- | --- | --- | --- | --- | --- | --- | --- | --- | --- | --- | --- | --- |
| ER %, Mean ± SEM |  | 80.9 ± 1.3 | 77.4 ± 1.7 | 73.3 ± 1.7 | **<0.001** | 77.9 ± 1.5 | 77.8 ± 1.6 | 75.9 ± 1.8 | 0.899 | 79.3 ± 1.4 | 74.4 ± 1.9 | 78.0 ± 1.4 | 0.252 |
| PR status, N (%) | Positive | 121 (70.8) | 121 (70.7) | 119 (69.6) | 0.953 | 123 (71.9) | 125 (73.1) | 113 (66.1) | 0.219 | 119 (69.6) | 123 (71.9) | 119 (69.6) | 0.868 |
|  | Negative | 49 (28.7) | 48 (28.1) | 49 (28.6) |  | 48 (28.1) | 44 (25.7) | 54 (31.6) |  | 51 (29.8) | 46 (26.9) | 49 (28.7) |  |
|  | Unknown | 1 (0.5) | 2 (1.2) | 3 (1.8) |  | 0 (0.0) | 2 (1.2) | 4 (2.3) |  | 1 (0.6) | 2 (1.2) | 3 (1.7) |  |
| Tumour size, N (%) | ≤20 | 137 (80.1) | 139 (81.3) | 141 (82.5) | 0.983 | 124 (72.5) | 140 (81.9) | 153 (89.5) | **<0.001** | 124 (72.5) | 143 (83.6) | 150 (87.7) | **<0.001** |
|  | >20 | 32 (18.7) | 30 (17.5) | 28 (16.3) |  | 47 (27.5) | 27 (15.8) | 16 (9.4) |  | 45 (26.3) | 28 (16.4) | 17 (9.9) |  |
|  | Unknown | 2 (1.2) | 2 (1.2) | 2 (1.2) |  | 0 (0.0) | 4 (2.3) | 2 (1.1) |  | 2 (1.2) | 0 (0.0) | 4 (2.4) |  |
| Tumour grade, N (%) | 1 | 37 (21.6) | 42 (24.6) | 36 (21.1) | 0.944 | 25 (14.6) | 33 (19.3) | 57 (33.3) | **<0.001** | 13 (7.6) | 36 (21.1) | 66 (38.6) | **<0.001** |
|  | 2 | 111 (64.9) | 104 (60.8) | 111 (64.8) |  | 107 (62.6) | 114 (66.7) | 105 (61.4) |  | 114 (66.7) | 112 (65.5) | 100 (58.5) |  |
|  | 3 | 21 (12.3) | 21 (12.3) | 22 (12.9) |  | 35 (20.5) | 23 (13.5) | 6 (3.5) |  | 41 (24.0) | 19 (11.1) | 4 (2.3) |  |
|  | Unknown | 2 (1.2) | 4 (2.3) | 2 (1.2) |  | 4 (2.3) | 1 (0.5) | 3 (1.8) |  | 3 (1.7) | 4 (2.3) | 1 (0.6) |  |
| Ki67 status, N (%) | Positive | 35 (20.5) | 29 (17.0) | 41 (24.0) | 0.488 | 47 (27.5) | 36 (21.1) | 22 (12.9) | **0.006** | 57 (33.4) | 32 (18.7) | 16 (9.4) | **<0.001** |
|  | Negative | 130 (76.0) | 133 (77.7) | 121 (70.7) |  | 120 (70.2) | 127 (74.3) | 137 (80.1) |  | 109 (63.7) | 130 (76.0) | 145 (84.8) |  |
|  | Unknown | 6 (3.5) | 9 (5.3) | 9 (5.3) |  | 4 (2.3) | 8 (4.6) | 12 (7.0) |  | 5 (2.9) | 9 (5.3) | 10 (5.8) |  |
| MINDACT clinical risk, N (%) | C-high | 36 (21.1) | 36 (21.1) | 40 (23.4) | 0.936 | 58 (33.9) | 38 (22.2) | 16 (9.4) | **<0.001** | 62 (36.3) | 32 (18.7) | 18 (10.5) | **<0.001** |
|  | C-low | 131 (76.6) | 130 (76.0) | 128 (74.8) |  | 109 (63.7) | 128 (74.9) | 152 (88.8) |  | 105 (61.4) | 135 (78.9) | 149 (87.1) |  |
|  | Unknown | 4 (2.3) | 5 (2.9) | 3 (1.8) |  | 4 (2.4) | 5 (2.9) | 3 (1.8) |  | 4 (2.3) | 4 (2.4) | 4 (2.4) |  |

<sup>a</sup>p-value calculated by Fisher's exact test for all variables except for the ER % and PR %. Kruskal-Wallis was used for those variables

### Long-term Tamoxifen benefit within TME tertiles

Next, we assessed tamoxifen benefit - defined as differences in distant recurrence-free interval (DRFI) between treated and untreated patients - across immune, endothelial, and fibroblast tertiles, as well as within individual immune cell type tertiles. Univariable Kaplan–Meier analyses demonstrated a statistically significant improvement in long-term DRFI in/among tamoxifen therapy-treated patients with lower relative immune score abundances but not intermediate or high, compared to patients who did not receive tamoxifen therapy (log-rank p < 0.001, for tamoxifen immune score low tertile vs. untreated patients, Figure 3A-C). Similarly, tamoxifen conferred a significant benefit in the endothelial intermediate tertile (log-rank p < 0.001, Figure 3E) and fibroblast low and intermediate tertiles (log-rank p = 0.042 and p = 0.009, respectively, Figure 3G-H) relative to patients who did not receive tamoxifen therapy. Kaplan–Meier analyses of distant recurrence by individual immune cell types revealed a similar trend to the aggregated immune score, whereby a statistically significant tamoxifen benefit was observed in the majority of immune cell low but not high tertiles (log-rank p <0.05 for all comparisons, Additional file 2A-P, adjusted for multiple testing using Bonferroni correction). Similar findings were seen in multivariable Cox proportional hazard modelling where tamoxifen conferred significantly improved long-term DRFI in tumours with lower immune cell abundance (adjusted Hazard Ratio (aHR) = 0.17; 95% CI, 0.08–0.40), intermediate endothelial cell abundance (aHR = 0.21; 95% CI, 0.09–0.51), and both low and intermediate fibroblast abundances (aHR = 0.50; 95% CI, 0.27–0.93, and aHR = 0.36; 95% CI, 0.17–0.77, respectively), compared to untreated patients (Figure 4). Similarly, individual immune cell types also revealed improved DRFI for tamoxifen-treated patients for all immune cell low tertiles (Additional file 3). A significant interaction between immune tertiles and tamoxifen treatment was also observed (p = 0.008, data not shown), indicating that the impact of immune infiltration differed by treatment group. No statistically significant interaction was found between tamoxifen treatment and endothelial or fibroblast tertiles (p = 0.850 and p = 0.800, respectively).

**Figure 3.**
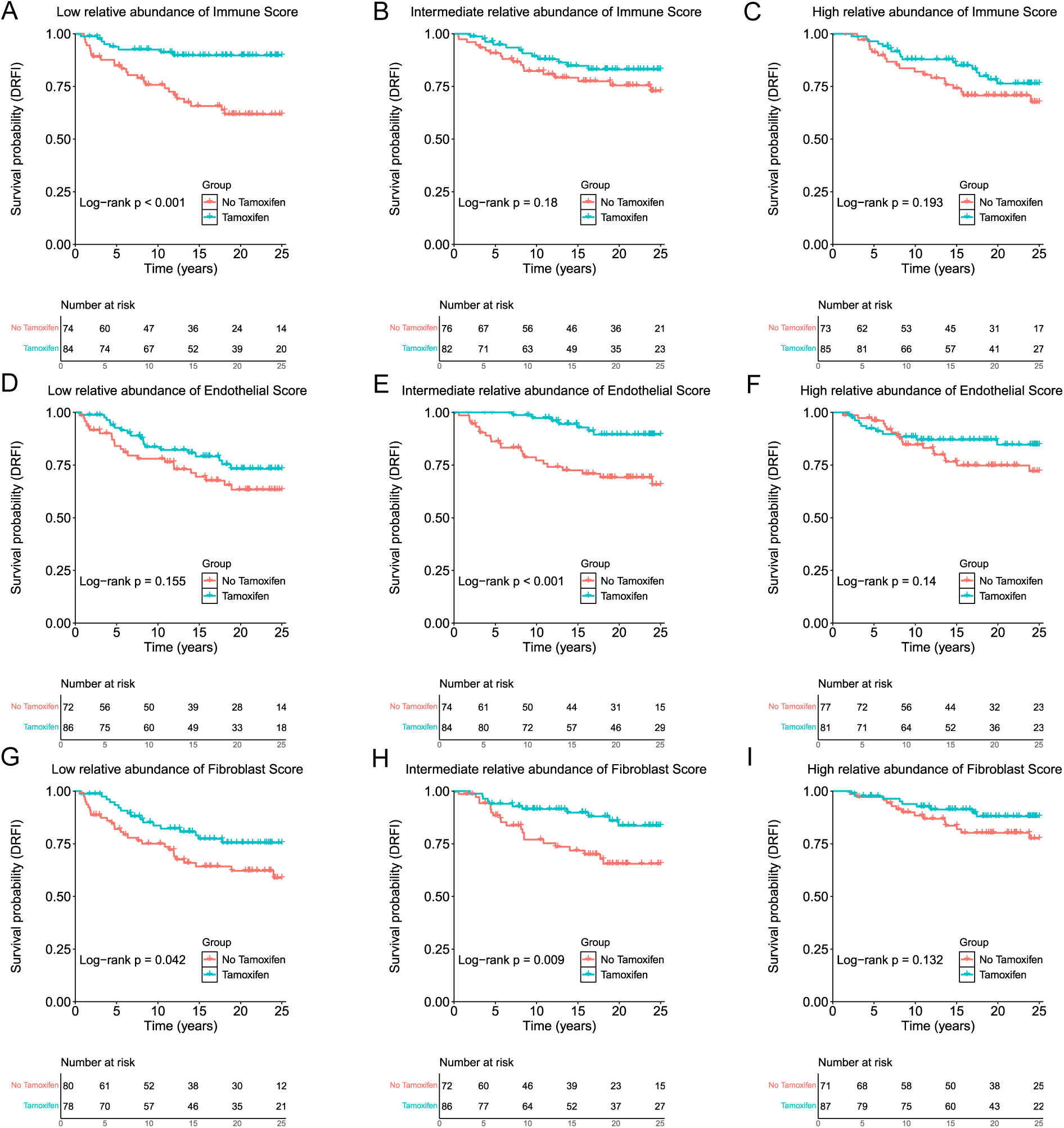
Univariable Kaplan-Meier analysis of DRFI in tamoxifen-treated patients compared to non-tamoxifen treated patients across tertile scores. Comparisons represent treatment benefit within each TME tertile (tamoxifen-treated vs. untreated). (A-C) Low, Intermediate and High relative abundance tertiles for the Immune score. (D-F) Low, Intermediate and High relative abundance tertiles for the Endothelial score. (G-I) Low, Intermediate and High relative abundance tertiles for the Fibroblast score.

**Figure 4.**
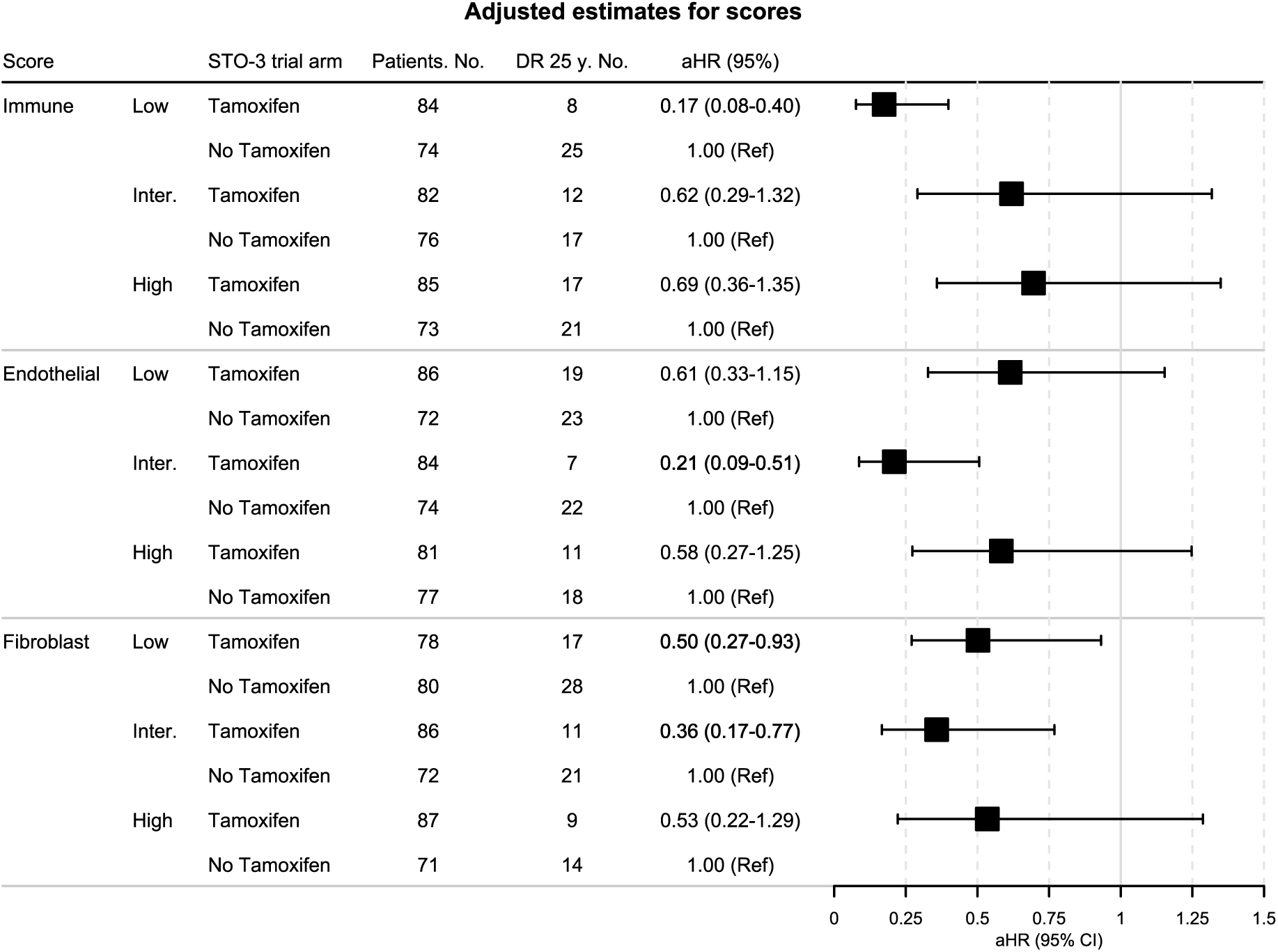
Multivariable Cox proportional hazard regression analysis of DRFI in tamoxifen-treated patients compared to untreated patients across tertile scores. Multivariable analyses were adjusted for clinical information and tumour characteristics, including age, tumour size, tumour grade, progesterone receptor status, and Ki-67 status, and the administration of radiotherapy. DRFI, distant recurrence-free interval; DR, distant recurrence; aHR, adjusted hazard ratio; Ref, reference.

### Distinct molecular profiles associated with tumour composition

In order to better understand why tamoxifen benefit is observed in specific TME tertiles we next performed differential expression with subsequent pathway enrichment analyses. When comparing low vs. high tertiles for immune and fibroblast scores we found that tumours with low scores for both cell types were enriched for early and late oestrogen response, MYC targets, and oxidative phosphorylation hallmark pathways, while interferon gamma response and allograft rejection were downregulated (Figure 5). Endothelial intermediate tumours were compared to low and high subgroups separately and MYC targets along with the oxidative phosphorylation were the only statistically significant downregulated pathways in both analyses (Additional file 4). Interestingly, hallmark oestrogen response early and late pathways were downregulated in the intermediate compared to low tertile for endothelial scores, although this difference was not significant when comparing intermediate to high. Finally, to ensure that differentially expressed genes were not the same as those that define the ConsensusTME cell type signatures we applied the Jaccard similarity index. No significant overlap was found between the immune-, fibroblast-, and endothelial-associated genes and the differentially expressed genes underlying their respective enriched pathways, suggesting distinct molecular processes for each pathway enrichment (data not shown).

**Figure 5.**
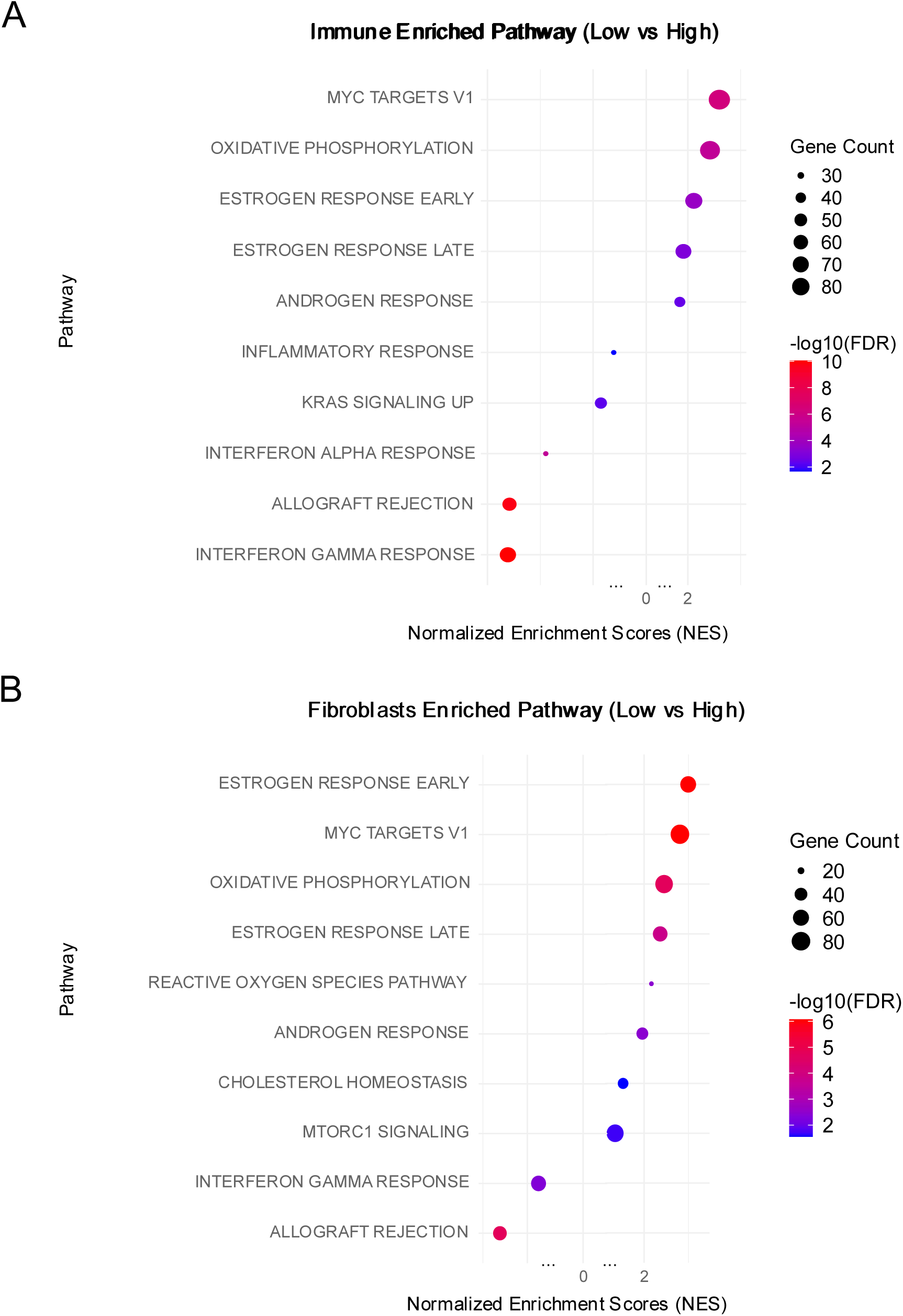
Dot plot representation of the most and least enriched pathways in low vs high for (A) immune score, and (B) fibroblast score. The circle’s size represents the number of genes in each pathway, and the colour of the qscore for each pathway.

## DISCUSSION

In this study, we examined the long-term benefit of tamoxifen treatment according to tumour microenvironment (TME) baseline composition in postmenopausal patients with ER+ HER2-breast cancer. Using the randomised design of the STO-3 trial including a control arm with no endocrine therapy, we were able to compare tamoxifen-treated patients versus untreated across tertiles of immune, endothelial, and fibroblast abundance. We found that low immune cell, intermediate endothelial cell, and low to intermediate fibroblast cell abundances are each associated with a greater long-term benefit from tamoxifen. Moreover, we identified differentially expressed hallmark pathways in these groups including the enrichment of oestrogen response, MYC targets and oxidative phosphorylation in immune and fibroblast low tertiles.

Comparisons with previous studies are challenging due to the unique long-term follow-up of our cohort and the direct clinical trial arm comparisons within immune and stromal abundance tertiles. Nevertheless, several studies align with our finding that low immune cell abundance may be linked to greater tamoxifen benefit in ER-positive breast cancer. Ali *et al.* used CIBERSORT to show that ER-positive tumours lacking immune infiltration had distinct biology and intermediate survival outcomes(8). They reported poorer survival with increased M0 macrophages and regulatory T cells on the ER+HER- group(8). Similarly, Sobral-Leite *et al.* observed that a higher abundance of CD4+ or CD8+ T-cells were associated with poorer prognosis in ER+ HER2- low grade breast cancer patients from a randomised clinical trial (23). Heindl *et al.* also found that high immune spatial scores (but not abundance scores) were associated with worse 10-year survival in the ATAC cohort (24). Collectively, these studies support our conclusion that immunologically low ER-positive HER2-negative tumours may have distinct biology associated with greater tamoxifen benefit.

Our work also identified a number of pathways enriched in tumours with low immune and fibroblast abundance (vs. high) and intermediate endothelial abundance (vs. low and high), some of which align with known biology for tamoxifen response. We found higher levels of oestrogen response pathways (in addition to a higher percentage of oestrogen positive cells) in tumours with a lower immune and fibroblast scores. This is likely directly related to the DRFI improvement found following tamoxifen treatment for these patient subgroups (Figures 3 and 4) and is in line with studies showing that tumours with a higher percentage of ER-positive cells tend to respond better to endocrine therapy(25,26) . Similarly, the higher levels of oxidative phosphorylation and lower levels of interferon gamma pathways found in the same tumours are in line with previous links to improved efficacy of tamoxifen(27,28). Contrastingly however, oxidative phosphorylation was depleted in tumours with intermediate endothelial cell abundance. Of note, only ∼25% of endothelial intermediate tumours were also immune low which may help to explain these opposing findings (data not shown). Interpretation of intermediate biomarker expression is difficult however and we would urge caution in over-interpreting this specific result until independently validated. A more general clinical interpretation of our findings could be that low immune and fibroblast content may reflect reduced stromal remodeling and fewer physical or signaling barriers to therapy(29).

The strengths of our study include the long-term clinical follow-up of 25 years, the clinical trial design that allows direct comparison to a control arm who did not receive tamoxifen and the comprehensive nature of our analysis examining 18 TME cell types in a single study. The limitations are as follows: First, clinical recommendations for treatment and disease management have evolved since the trial’s commencement. The trial was conducted prior to the recommendation of aromatase inhibitors for postmenopausal patients instead of tamoxifen. Additionally, the treatment duration was shorter (2 years) and administered a higher dose tamoxifen dose than current guidelines. Second, whilst we analysed tumours from 513 patients in total, numbers are lower in subgroup tertile analyses with between approximately 70-85 patients in each arm. Reassuringly, however, our findings for individual immune cell types closely match those in our aggregated immune score (Additional file 2 and 3). Third, a general limitation to deconvolution methods is that they tend to show better correlation to fluorescence-activated cell sorting (FACS) when measuring cell proportions for the major immune cell types such as CD4+ T cells, CD8+ T cells, B cells, NK cells, and macrophages. Correlations to rarer immune cells e.g. regulatory T cells or dendritic cells tend to be poorer and, as such, less reliable(30). Fourth and relatedly, we are assessing the relative abundance of immune cells using bulk tumour transcriptomic analysis rather than whole section immunohistochemical slide staining. However, staining and scoring of 18 immune cell types using reliable markers is beyond the scope of the current project. Interestingly, advances in the ability of Artificial Intelligence (AI) algorithms to accurately estimate biomarker expression from whole section hematoxylin and eosin (H&E) slides may render this more feasible in near future(31,32). In line with these limitations, future studies should assess whether baseline TME composition also predicts benefit from extended tamoxifen therapy beyond 5 years. Moreover, validation in larger, independent cohorts, along with spatial and single-cell profiling, will be key to refining the role of the tumour microenvironment in guiding endocrine treatment decisions.

## CONCLUSION

Findings from the STO-3 trial with 25 years of follow-up show that baseline tumour microenvironment composition, particularly low immune, intermediate endothelial, and low to intermediate fibroblast abundance, has predictive value for tamoxifen benefit in ER+HER2-postmenopausal breast cancer. As one of the first studies to assess tamoxifen benefit relative to TME in a randomised, no-treatment control setting, our findings highlight the importance of microenvironmental context in guiding endocrine therapy decisions for ER+HER2– breast cancer patients, a group historically considered immunologically cold.

## Data Availability

The data underlying this article cannot be shared publicly due to the privacy of the individuals who participated in the study.
All code used for the analyses presented in this manuscript is publicly available at https://github.com/pcr08/STO3_25y_TME_Tamoxifen

## ACKNOWLEDGMENTS

The authors express their gratitude to all the patients who took part in the STO trials, particularly STO-3, and to the Stockholm Breast Cancer Study Group for making this study possible. The deconvolution computations were enabled by resources provided by the National Academic Infrastructure for Supercomputing in Sweden (NAISS), partially funded by the Swedish Research Council through grant agreement no. 2022-06725.

## Abbreviations

AI: Artificial Intelligence
ATAC: Arimidex, Tamoxifen, Alone or in Combination
BRCA: Breast Cancer
DRFI: Distant Recurrence-Free Interval
ER: Oestrogen Receptor
ESMO: European Society for Medical Oncology
FFPE: Formalin-Fixed Paraffin-Embedded
HR: Hormone receptor
HER2: Human Epidermal growth factor 2
H&E: Hematoxylin and Eosin
IHC: Immunohistochemistry
NES: Normalized Enriched Scores
NK: Natural Killer
NO: Nitric Oxide
PR: Progesterone receptor
RNA: Ribonucleic acid
SERM: Selective Oestrogen Receptor Modulators
STO-3: Stockholm tamoxifen randomised trial 3
TAMs: Tumour-Associated Macrophages
TIL: Tumour-infiltrating lymphocytes
TME: Tumour Microenvironment
TNBC: Triple Negative Breast Cancer
UPGMA: Unweighted Pair Group Method with Arithmetic Mean

## References

1. Siegel RL, Kratzer TB, Giaquinto AN, Sung H, Jemal A. Cancer statistics, 2025. CA Cancer J Clin. 2025 Jan;75(1):10–45.

2. Arnold M, Morgan E, Rumgay H, Mafra A, Singh D, Laversanne M, et al. Current and future burden of breast cancer: Global statistics for 2020 and 2040. The Breast. 2022 Dec;66:15–23.

3. Guo L, Kong D, Liu J, Zhan L, Luo L, Zheng W, et al. Breast cancer heterogeneity and its implication in personalized precision therapy. Experimental Hematology & Oncology 2023 12:1 [Internet]. 2023 Jan 9 [cited 2024 Feb 27];12(1):1–27. Available from: https://ehoonline.biomedcentral.com/articles/10.1186/s40164-022-00363-1

4. Zhao Y, Shen M, Wu L, Yang H, Yao Y, Yang Q, et al. Stromal cells in the tumor microenvironment: accomplices of tumor progression? Vol. 14, Cell Death and Disease. Springer Nature; 2023.

5. Deepak KGK, Vempati R, Nagaraju GP, Dasari VR, Nagini S, Rao DN, et al. Tumor microenvironment: Challenges and opportunities in targeting metastasis of triple negative breast cancer. Pharmacol Res. 2020 Mar 1;153:104683.

6. Zhao H, Yin X, Wang L, Liu K, Liu W, Bo L, et al. Identifying tumour microenvironment-related signature that correlates with prognosis and immunotherapy response in breast cancer. Scientific Data 2023 10:1 [Internet]. 2023 Mar 3 [cited 2024 Feb 27];10(1):1–14. Available from: https://www.nature.com/articles/s41597-023-02032-2

7. Segovia-Mendoza M, Morales-Montor J. Immune tumor microenvironment in breast cancer and the participation of estrogens and its receptors into cancer physiopathology. Vol. 10, Frontiers in Immunology. Frontiers Media S.A.; 2019.

8. Ali HR, Chlon L, Pharoah PDP, Markowetz F, Caldas C. Patterns of Immune Infiltration in Breast Cancer and Their Clinical Implications: A Gene-Expression-Based Retrospective Study. PLoS Med. 2016 Dec 1;13(12).

9. Mahmoud SMA, Paish EC, Powe DG, Macmillan RD, Grainge MJ, Lee AHS, et al. Tumor-infiltrating CD8+ lymphocytes predict clinical outcome in breast cancer. Journal of Clinical Oncology. 2011 May 20;29(15):1949–55.

10. Egelston CA, Guo W, Tan J, Avalos C, Simons DL, Lim MH, et al. Tumor-infiltrating exhausted CD8+ T cells dictate reduced survival in premenopausal estrogen receptor–positive breast cancer. JCI Insight. 2022 Feb 8;7(3).

11. Oshi M, Asaoka M, Tokumaru Y, Yan L, Matsuyama R, Ishikawa T, et al. CD8 T Cell Score as a Prognostic Biomarker for Triple Negative Breast Cancer. Int J Mol Sci [Internet]. 2020 Sep 2 [cited 2024 Feb 27];21(18):1–16. Available from: /pmc/articles/PMC7555570/

12. Rutqvist LE, Johansson H, Group) (on behalf of the Stockholm Breast Cancer Study. Long-term follow-up of the randomized Stockholm trial on adjuvant tamoxifen among postmenopausal patients with early stage breast cancer. Acta Oncol (Madr) [Internet]. 2007 [cited 2024 Mar 22];46(2):133–45. Available from: https://www.tandfonline.com/doi/abs/10.1080/02841860601034834

13. Lindström LS, Yau C, Czene K, Thompson CK, Hoadley KA, Van’t Veer LJ, et al. Intratumor Heterogeneity of the Estrogen Receptor and the Long-term Risk of Fatal Breast Cancer. JNCI: Journal of the National Cancer Institute [Internet]. 2018 Jul 1 [cited 2024 Mar 22];110(7):726–33. Available from: 10.1093/jnci/djx270

14. Johansson A, Yu NY, Iftimi A, Tobin NP, van ’t Veer L, Nordenskjöld B, et al. Clinical and molecular characteristics of estrogen receptor-positive ultralow risk breast cancer tumors identified by the 70-gene signature. Int J Cancer [Internet]. 2022 Jun 15 [cited 2024 Mar 22];150(12):2072–82. Available from: https://onlinelibrary.wiley.com/doi/full/10.1002/ijc.33969

15. Barlow L, Westergren K, Holmberg L, Tälback M. The completeness of the Swedish Cancer Register - A sample survey for year 1998. Acta Oncol (Madr). 2009 Jan;48(1):27–33.

16. Brooke HL, Talbäck M, Hörnblad J, Johansson LA, Ludvigsson JF, Druid H, et al. The Swedish cause of death register. Eur J Epidemiol. 2017 Sep 1;32(9):765–73.

17. Emilsson L, Lindahl B, Köster M, Lambe M, Ludvigsson JF. Review of 103 Swedish Healthcare Quality Registries. J Intern Med. 2015 Jan 1;277(1):94–136.

18. Svenska bröstcancergruppen (Swedish Breast Cancer Group). Nationellt Vardprogram Bröstcancer (Swedish_National_Guidelines). 2019;

19. Jerevall PL, Ma XJ, Li H, Salunga R, Kesty NC, Erlander MG, et al. Prognostic utility of HOXB13: IL17BR and molecular grade index in early-stage breast cancer patients from the Stockholm trial. Br J Cancer. 2011 May 24;104(11):1762–9.

20. Jimenez-Sanchez A, Cast O, Miller ML. Comprehensive benchmarking and integration of tumor microenvironment cell estimation methods. Cancer Res [Internet]. 2019 Dec 15 [cited 2024 Mar 22];79(24):6238–46. Available from: /cancerres/article/79/24/6238/639705/Comprehensive-Benchmarking-and-Integration-of

21. Hudis CA, Barlow WE, Costantino JP, Gray RJ, Pritchard KI, Chapman JAW, et al. Proposal for standardized definitions for efficacy end points in adjuvant breast cancer trials: The STEEP system. Journal of Clinical Oncology. 2007 May 20;25(15):2127–32.

22. Cardoso F, Van’t Veer L, Rutgers E, Loi S, Mook S, Piccart-Gebhart MJ. Clinical application of the 70-gene profile: The MINDACT trial. Vol. 26, Journal of Clinical Oncology. 2008. p. 729–35.

23. Sobral-Leite M, Salomon I, Opdam M, Kruger DT, Beelen KJ, Van Der Noort V, et al. Cancer-immune interactions in ER-positive breast cancers: PI3K pathway alterations and tumor-infiltrating lymphocytes. Breast Cancer Research. 2019 Aug 7;21(1).

24. Heindl A, Sestak I, Naidoo K, Cuzick J, Dowsett M, Yuan Y. Relevance of Spatial Heterogeneity of Immune Infiltration for Predicting Risk of Recurrence after Endocrine Therapy of ER+ Breast Cancer. J Natl Cancer Inst. 2018 Feb 1;110(2):166–75.

25. Ellis MJ, Ma C. Letrozole in the neoadjuvant setting: The P024 trial. Vol. 105, Breast Cancer Research and Treatment. Kluwer Academic Publishers; 2007. p. 33–43.

26. Makhlouf S, Quinn C, Toss M, Alsaleem M, Atallah NM, Ibrahim A, et al. Quantitative expression of oestrogen receptor in breast cancer: Clinical and molecular significance. Eur J Cancer. 2024 Jan 1;197.

27. Bekele RT, Venkatraman G, Liu RZ, Tang X, Mi S, Benesch MGK, et al. Oxidative stress contributes to the tamoxifen-induced killing of breast cancer cells: Implications for tamoxifen therapy and resistance. Sci Rep. 2016 Feb 17;6.

28. Post AEM, Smid M, Nagelkerke A, Martens JWM, Bussink J, Sweep FCGJ, et al. Interferon-stimulated genes are involved in cross-resistance to radiotherapy in tamoxifen-resistant breast cancer. Clinical Cancer Research. 2018 Jul 15;24(14):3397–408.

29. Kalluri R. The biology and function of fibroblasts in cancer. Vol. 16, Nature Reviews Cancer. Nature Publishing Group; 2016. p. 582–98.

30. Avila Cobos F, Alquicira-Hernandez J, Powell JE, Mestdagh P, De Preter K. Benchmarking of cell type deconvolution pipelines for transcriptomics data. Nat Commun. 2020 Dec 1;11(1).

31. Arslan S, Schmidt J, Bass C, Mehrotra D, Geraldes A, Singhal S, et al. A systematic pan-cancer study on deep learning-based prediction of multi-omic biomarkers from routine pathology images. Communications Medicine. 2024 Dec 1;4(1).

32. Shamai G, Binenbaum Y, Slossberg R, Duek I, Gil Z, Kimmel R. Artificial Intelligence Algorithms to Assess Hormonal Status from Tissue Microarrays in Patients with Breast Cancer. JAMA Netw Open. 2019 Jul 26;2(7).

