## Supplemental Figure 1 for "The tumour microenvironment influences long-term tamoxifen benefit in postmenopausal ER+/HER2- breast cancer patients"

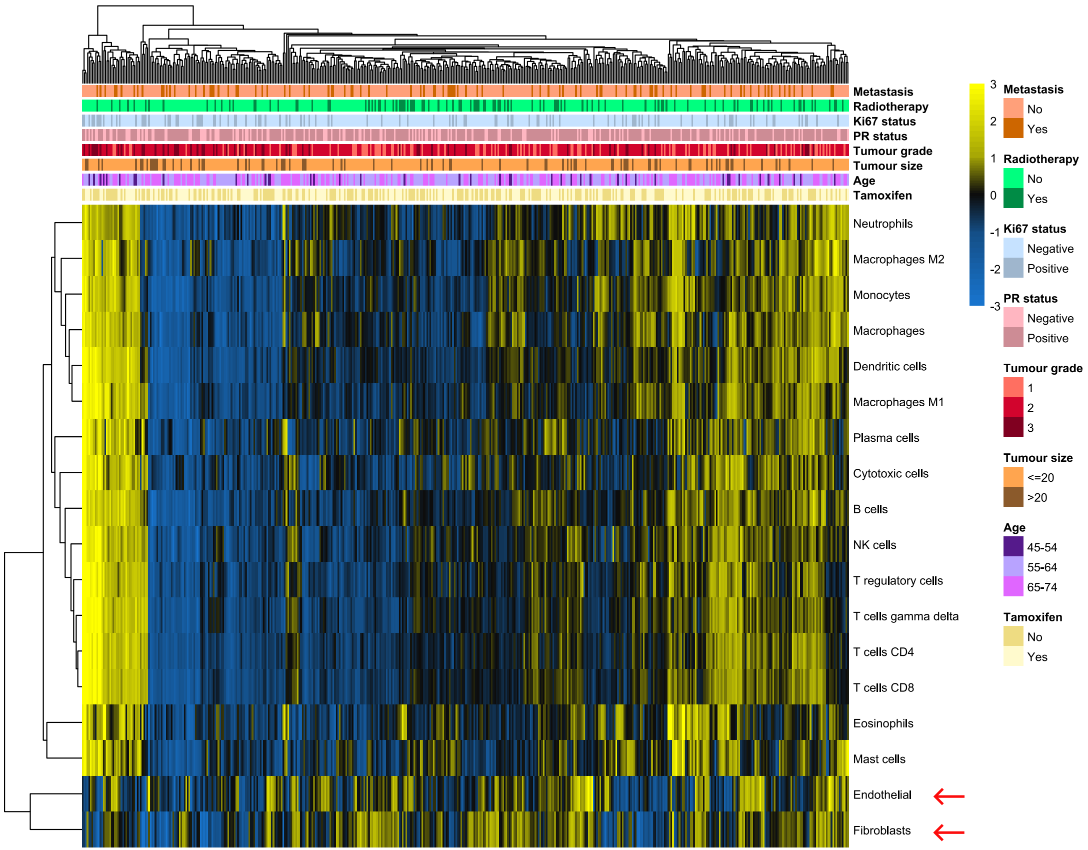


**Supplementary Figure S1**: Unsupervised heatmap displaying hierarchical clustering of TME cell types across patient samples. The colour gradient represents scaled values, with blue indicating low and yellow high relative abundance. Annotations above the heatmap denote clinical and molecular features, including Metastasis, Radiotherapy, Ki-67 status, PR status, Tumour grade, Age, and Tamoxifen treatment. Rows correspond to immune cell types, and columns to patients.
