## Supplemental Figure 2 for "The tumour microenvironment influences long-term tamoxifen benefit in postmenopausal ER+/HER2- breast cancer patients"

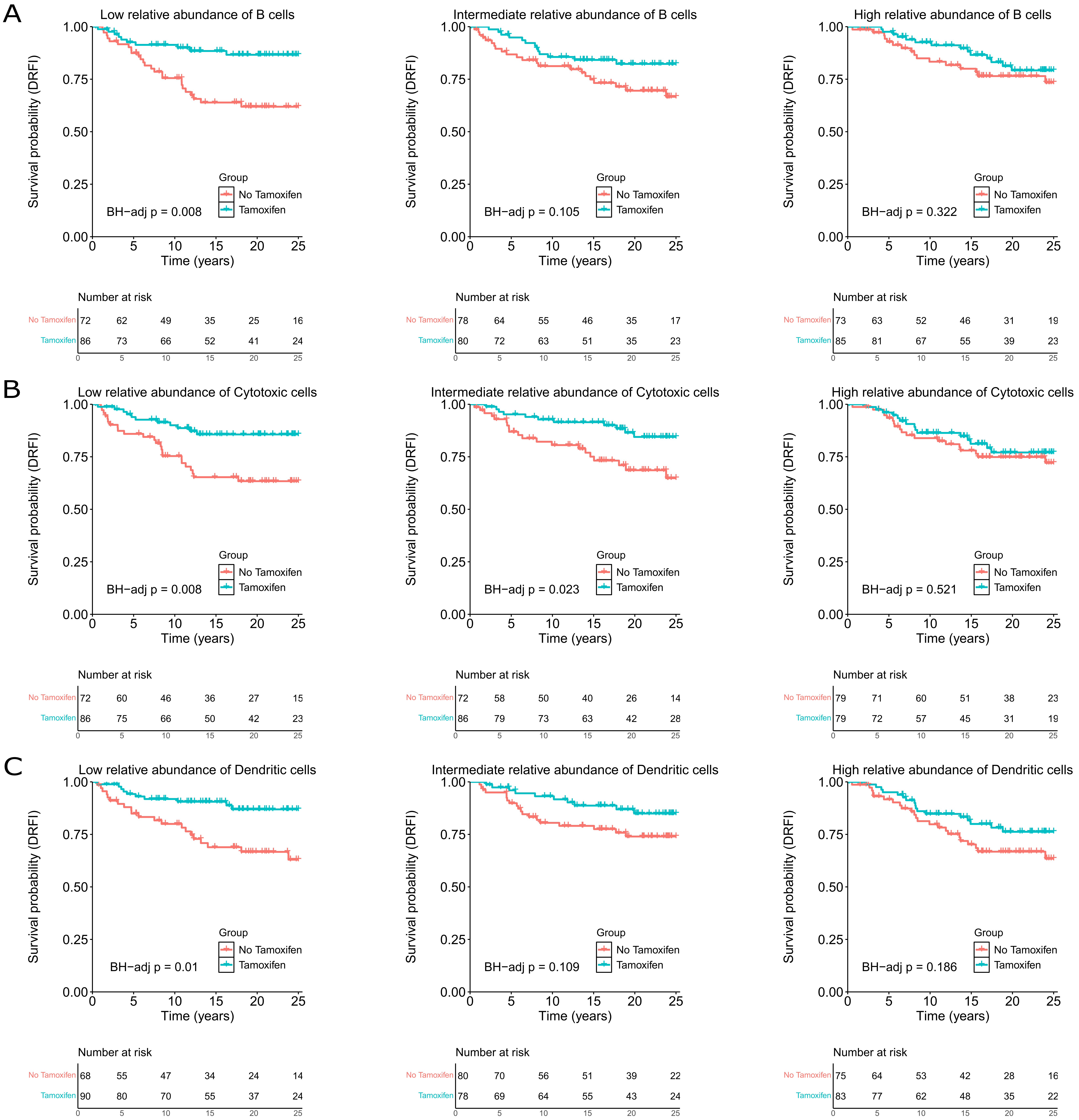
A
Low relative abundance of B cells
Survival probability (DRFI)
Time (years)
Group
No Tamoxifen
Tamoxifen
BH−adj p = 0.008
Number at risk
No Tamoxifen 72 62 49 35 25 16
Tamoxifen 86 73 66 52 41 24
Intermediate relative abundance of B cells
BH−adj p = 0.105
No Tamoxifen 78 64 55 46 35 17
Tamoxifen 80 72 63 51 35 23
High relative abundance of B cells
BH−adj p = 0.322
No Tamoxifen 73 63 52 46 31 19
Tamoxifen 85 81 67 55 39 23
B
Low relative abundance of Cytotoxic cells
BH−adj p = 0.008
No Tamoxifen 72 60 46 36 27 15
Tamoxifen 86 75 66 50 42 23
Intermediate relative abundance of Cytotoxic cells
BH−adj p = 0.023
No Tamoxifen 72 58 50 40 26 14
Tamoxifen 86 79 73 63 42 28
High relative abundance of Cytotoxic cells
BH−adj p = 0.521
No Tamoxifen 79 71 60 51 38 23
Tamoxifen 79 72 57 45 31 19
C
Low relative abundance of Dendritic cells
BH−adj p = 0.01
No Tamoxifen 68 55 47 34 24 14
Tamoxifen 90 80 70 55 37 24
Intermediate relative abundance of Dendritic cells
BH−adj p = 0.109
No Tamoxifen 80 70 56 51 39 22
Tamoxifen 78 69 64 55 43 24
High relative abundance of Dendritic cells
BH−adj p = 0.186
No Tamoxifen 75 64 53 42 28 16
Tamoxifen 83 77 62 48 35 22

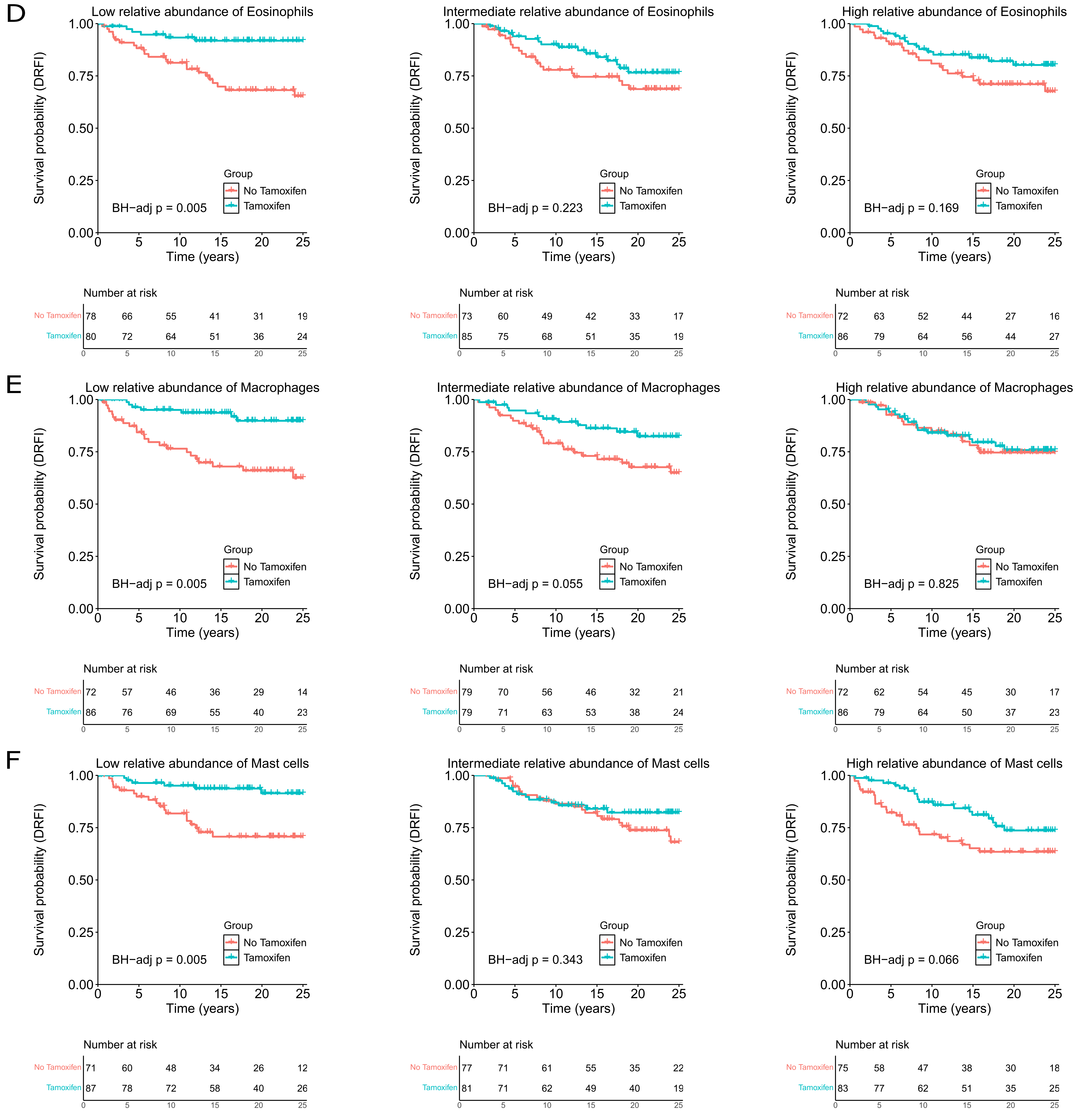
D
Low relative abundance of Eosinophils
Intermediate relative abundance of Eosinophils
High relative abundance of Eosinophils
Survival probability (DRFI)
1.00
0.75
0.50
0.25
0.00
0
5
10
15
20
25
Time (years)
Group
No Tamoxifen
Tamoxifen
BH−adj p = 0.005
BH−adj p = 0.223
BH−adj p = 0.169
Number at risk
No Tamoxifen 78 66 55 41 31 19
Tamoxifen 80 72 64 51 36 24
No Tamoxifen 73 60 49 42 33 17
Tamoxifen 85 75 68 51 35 19
No Tamoxifen 72 63 52 44 27 16
Tamoxifen 86 79 64 56 44 27
E
Low relative abundance of Macrophages
Intermediate relative abundance of Macrophages
High relative abundance of Macrophages
BH−adj p = 0.005
BH−adj p = 0.055
BH−adj p = 0.825
Number at risk
No Tamoxifen 72 57 46 36 29 14
Tamoxifen 86 76 69 55 40 23
No Tamoxifen 79 70 56 46 32 21
Tamoxifen 79 71 63 53 38 24
No Tamoxifen 72 62 54 45 30 17
Tamoxifen 86 79 64 50 37 23
F
Low relative abundance of Mast cells
Intermediate relative abundance of Mast cells
High relative abundance of Mast cells
BH−adj p = 0.005
BH−adj p = 0.343
BH−adj p = 0.066
Number at risk
No Tamoxifen 71 60 48 34 26 12
Tamoxifen 87 78 72 58 40 26
No Tamoxifen 77 71 61 55 35 22
Tamoxifen 81 71 62 49 40 19
No Tamoxifen 75 58 47 38 30 18
Tamoxifen 83 77 62 51 35 25

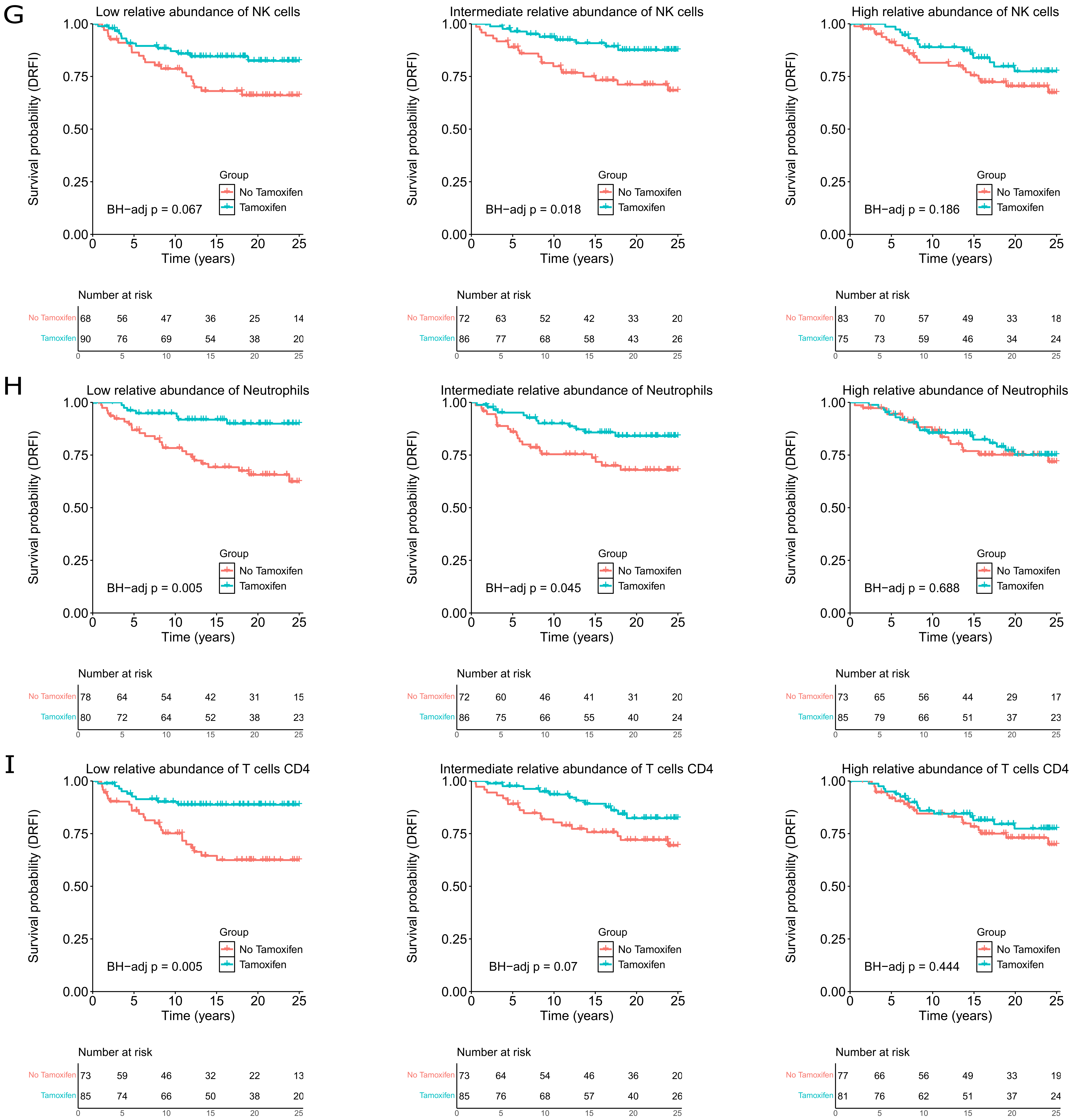
G
Low relative abundance of NK cells
BH−adj p = 0.067
Intermediate relative abundance of NK cells
BH−adj p = 0.018
High relative abundance of NK cells
BH−adj p = 0.186
Survival probability (DRFI)
Time (years)
Group
No Tamoxifen
Tamoxifen
Number at risk
No Tamoxifen 68 56 47 36 25 14
Tamoxifen 90 76 69 54 38 20
No Tamoxifen 72 63 52 42 33 20
Tamoxifen 86 77 68 58 43 26
No Tamoxifen 83 70 57 49 33 18
Tamoxifen 75 73 59 46 34 24
H
Low relative abundance of Neutrophils
BH−adj p = 0.005
Intermediate relative abundance of Neutrophils
BH−adj p = 0.045
High relative abundance of Neutrophils
BH−adj p = 0.688
No Tamoxifen 78 64 54 42 31 15
Tamoxifen 80 72 64 52 38 23
No Tamoxifen 72 60 46 41 31 20
Tamoxifen 86 75 66 55 40 24
No Tamoxifen 73 65 56 44 29 17
Tamoxifen 85 79 66 51 37 23
I
Low relative abundance of T cells CD4
BH−adj p = 0.005
Intermediate relative abundance of T cells CD4
BH−adj p = 0.07
High relative abundance of T cells CD4
BH−adj p = 0.444
No Tamoxifen 73 59 46 32 22 13
Tamoxifen 85 74 66 50 38 20
No Tamoxifen 73 64 54 46 36 20
Tamoxifen 85 76 68 57 40 26
No Tamoxifen 77 66 56 49 33 19
Tamoxifen 81 76 62 51 37 24

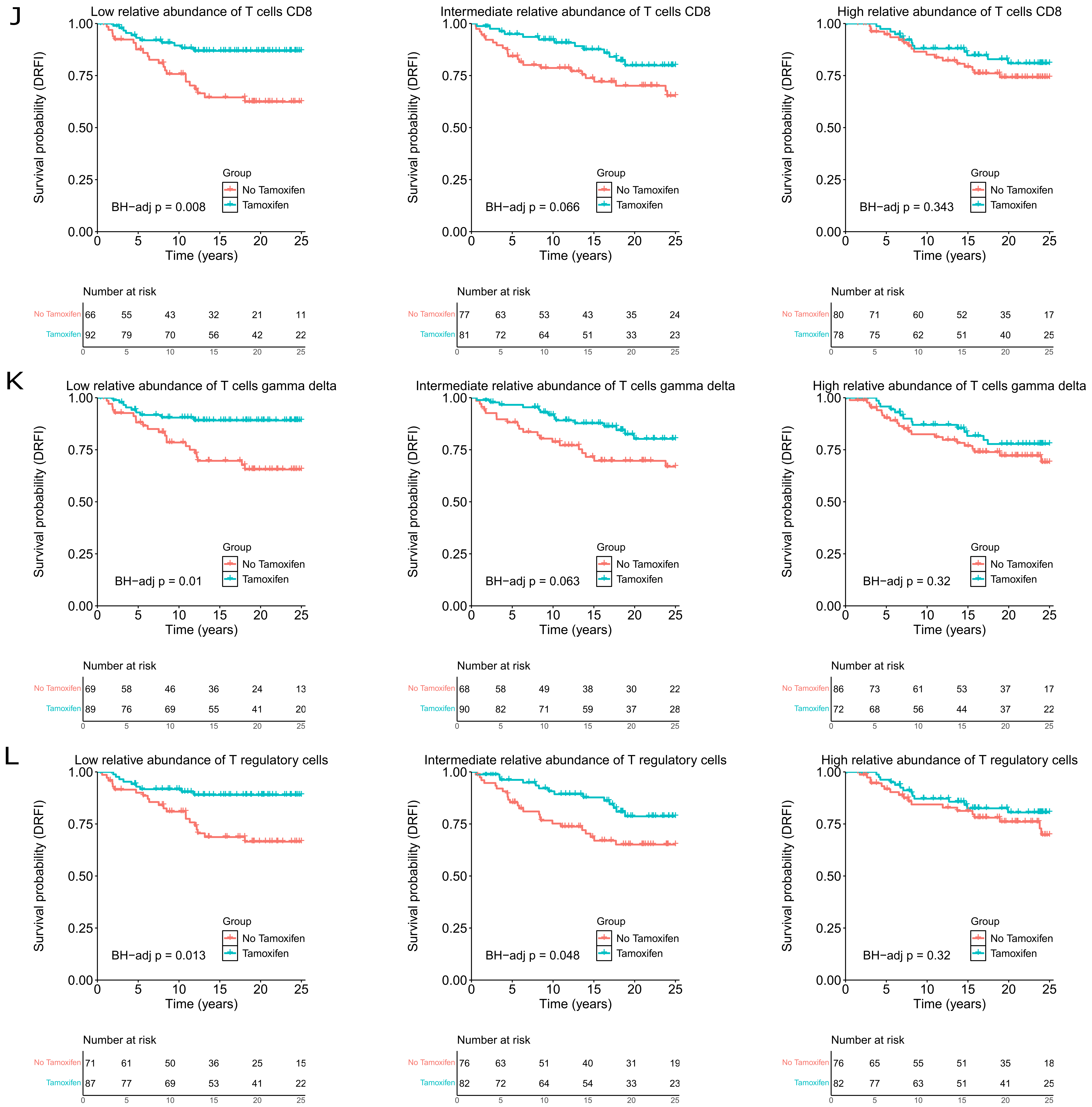
J
Low relative abundance of T cells CD8
Intermediate relative abundance of T cells CD8
High relative abundance of T cells CD8
Survival probability (DRFI)
Time (years)
Group
No Tamoxifen
Tamoxifen
BH−adj p = 0.008
BH−adj p = 0.066
BH−adj p = 0.343
Number at risk
No Tamoxifen 66 55 43 32 21 11
Tamoxifen 92 79 70 56 42 22
No Tamoxifen 77 63 53 43 35 24
Tamoxifen 81 72 64 51 33 23
No Tamoxifen 80 71 60 52 35 17
Tamoxifen 78 75 62 51 40 25
K
Low relative abundance of T cells gamma delta
Intermediate relative abundance of T cells gamma delta
High relative abundance of T cells gamma delta
BH−adj p = 0.01
BH−adj p = 0.063
BH−adj p = 0.32
No Tamoxifen 69 58 46 36 24 13
Tamoxifen 89 76 69 55 41 20
No Tamoxifen 68 58 49 38 30 22
Tamoxifen 90 82 71 59 37 28
No Tamoxifen 86 73 61 53 37 17
Tamoxifen 72 68 56 44 37 22
L
Low relative abundance of T regulatory cells
Intermediate relative abundance of T regulatory cells
High relative abundance of T regulatory cells
BH−adj p = 0.013
BH−adj p = 0.048
BH−adj p = 0.32
No Tamoxifen 71 61 50 36 25 15
Tamoxifen 87 77 69 53 41 22
No Tamoxifen 76 63 51 40 31 19
Tamoxifen 82 72 64 54 33 23
No Tamoxifen 76 65 55 51 35 18
Tamoxifen 82 77 63 51 41 25

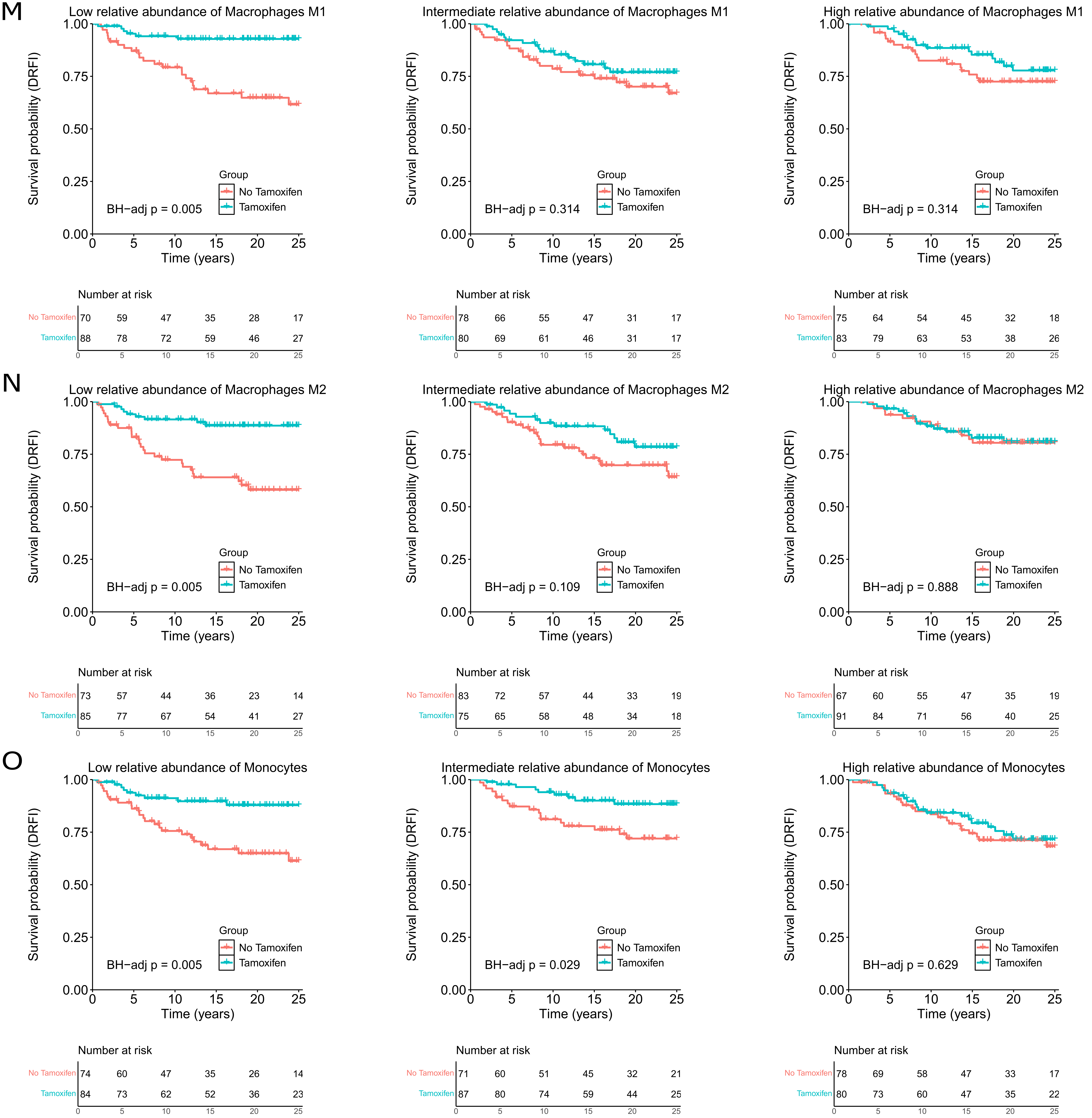
M
Low relative abundance of Macrophages M1
BH−adj p = 0.005
Intermediate relative abundance of Macrophages M1
BH−adj p = 0.314
High relative abundance of Macrophages M1
BH−adj p = 0.314
No Tamoxifen 70 59 47 35 28 17
Tamoxifen 88 78 72 59 46 27
No Tamoxifen 78 66 55 47 31 17
Tamoxifen 80 69 61 46 31 17
No Tamoxifen 75 64 54 45 32 18
Tamoxifen 83 79 63 53 38 26
N
Low relative abundance of Macrophages M2
BH−adj p = 0.005
Intermediate relative abundance of Macrophages M2
BH−adj p = 0.109
High relative abundance of Macrophages M2
BH−adj p = 0.888
No Tamoxifen 73 57 44 36 23 14
Tamoxifen 85 77 67 54 41 27
No Tamoxifen 83 72 57 44 33 19
Tamoxifen 75 65 58 48 34 18
No Tamoxifen 67 60 55 47 35 19
Tamoxifen 91 84 71 56 40 25
O
Low relative abundance of Monocytes
BH−adj p = 0.005
Intermediate relative abundance of Monocytes
BH−adj p = 0.029
High relative abundance of Monocytes
BH−adj p = 0.629
No Tamoxifen 74 60 47 35 26 14
Tamoxifen 84 73 62 52 36 23
No Tamoxifen 71 60 51 45 32 21
Tamoxifen 87 80 74 59 44 25
No Tamoxifen 78 69 58 47 33 17
Tamoxifen 80 73 60 47 35 22
Survival probability (DRFI)
Time (years)
Group
No Tamoxifen
Tamoxifen
Number at risk

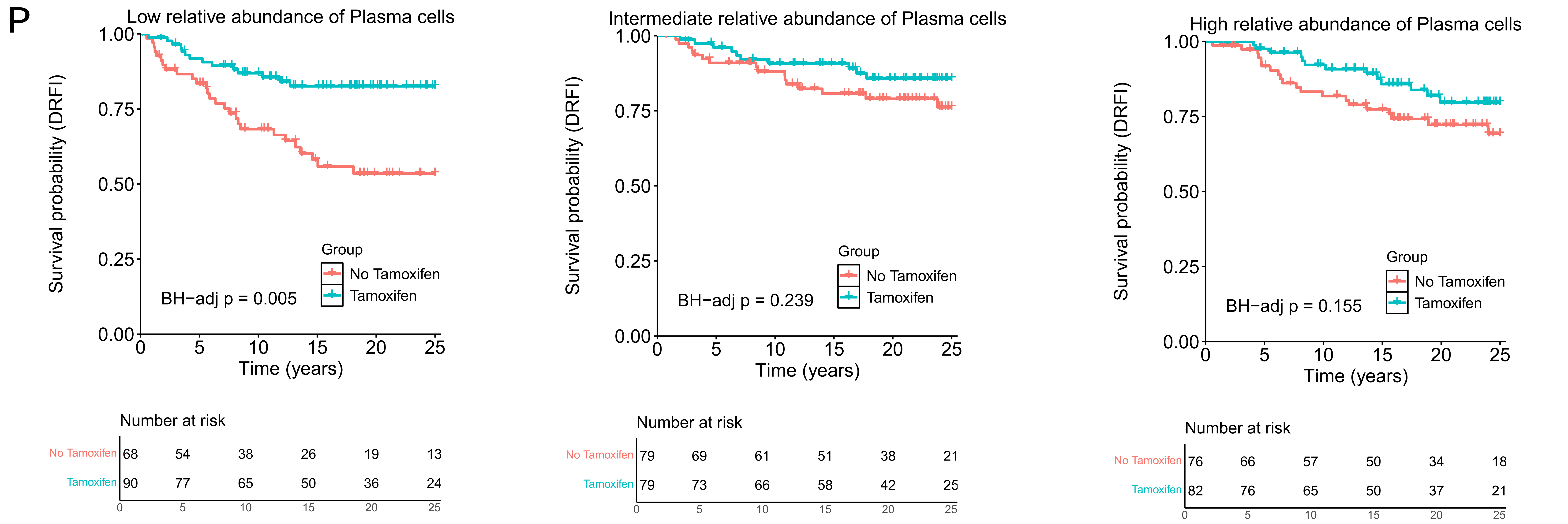


**Supplementary Figure S2:** Univariable Kaplan-Meier analysis of DRFI in tamoxifen-treated patients compared to no tamoxifen treated patients across tertile scores. Letters from A to Prepresent Low, Intermediate and High relative abundance tertiles for a specific celltype of the tumour microenvironment.
