## Supplemental Figure 3 for "The tumour microenvironment influences long-term tamoxifen benefit in postmenopausal ER+/HER2- breast cancer patients"

**Adjusted estimates for cell types**

| Cell type | | STO-3 trial arm | Patients, No. | DR 25 y. No. | aHR (95%) | BH-adj p |
|---|---|---|---|---|---|---|
| B cells | Low | Tamoxifen | 86 | 10 | **0.28 (0.13-0.60)** | **0.004** |
| | | No Tamoxifen | 72 | 25 | 1 (ref.) | |
| | Inter. | Tamoxifen | 80 | 13 | 0.45 (0.22-0.91) | 0.055 |
| | | No Tamoxifen | 78 | 22 | 1 (ref.) | |
| | High | Tamoxifen | 85 | 14 | 0.78 (0.37-1.68) | 0.604 |
| | | No Tamoxifen | 73 | 16 | 1 (ref.) | |
| Cytotoxic cells | Low | Tamoxifen | 86 | 11 | **0.30 (0.14-0.63)** | **0.005** |
| | | No Tamoxifen | 72 | 24 | 1 (ref.) | |
| | Inter. | Tamoxifen | 86 | 11 | **0.32 (0.15-0.70)** | **0.012** |
| | | No Tamoxifen | 72 | 20 | 1 (ref.) | |
| | High | Tamoxifen | 79 | 15 | 0.84 (0.42-1.69) | 0.679 |
| | | No Tamoxifen | 79 | 19 | 1 (ref.) | |
| Dendritic cells | Low | Tamoxifen | 90 | 10 | **0.24 (0.11-0.55)** | **0.003** |
| | | No Tamoxifen | 68 | 21 | 1 (ref.) | |
| | Inter. | Tamoxifen | 78 | 10 | 0.46 (0.21-1.01) | 0.100 |
| | | No Tamoxifen | 80 | 19 | 1 (ref.) | |
| | High | Tamoxifen | 83 | 17 | 0.60 (0.31-1.13) | 0.164 |
| | | No Tamoxifen | 75 | 23 | 1 (ref.) | |
| Eosinophils | Low | Tamoxifen | 80 | 6 | **0.14 (0.05-0.36)** | **0.001** |
| | | No Tamoxifen | 78 | 23 | 1 (ref.) | |
| | Inter. | Tamoxifen | 85 | 16 | 0.62 (0.32-1.23) | 0.235 |
| | | No Tamoxifen | 73 | 20 | 1 (ref.) | |
| | High | Tamoxifen | 86 | 15 | 0.55 (0.27-1.12) | 0.154 |
| | | No Tamoxifen | 72 | 20 | 1 (ref.) | |
| Macrophages | Low | Tamoxifen | 86 | 7 | **0.18 (0.08-0.43)** | **0.001** |
| | | No Tamoxifen | 72 | 23 | 1 (ref.) | |
| | Inter. | Tamoxifen | 79 | 12 | **0.40 (0.20-0.83)** | **0.030** |
| | | No Tamoxifen | 79 | 24 | 1 (ref.) | |
| | High | Tamoxifen | 86 | 18 | 1.09 (0.54-2.22) | 0.834 |
| | | No Tamoxifen | 72 | 16 | 1 (ref.) | |
| Mast cells | Low | Tamoxifen | 87 | 6 | **0.16 (0.06-0.43)** | **0.001** |
| | | No Tamoxifen | 71 | 18 | 1 (ref.) | |
| | Inter. | Tamoxifen | 81 | 13 | 0.70 (0.35-1.44) | 0.424 |
| | | No Tamoxifen | 77 | 20 | 1 (ref.) | |
| | High | Tamoxifen | 83 | 18 | 0.55 (0.30-1.03) | 0.107 |
| | | No Tamoxifen | 75 | 25 | 1 (ref.) | |
| NK cells | Low | Tamoxifen | 90 | 14 | **0.39 (0.19-0.80)** | **0.024** |
| | | No Tamoxifen | 68 | 21 | 1 (ref.) | |
| | Inter. | Tamoxifen | 86 | 9 | 0.39 (0.17-0.88) | 0.052 |
| | | No Tamoxifen | 72 | 20 | 1 (ref.) | |
| | High | Tamoxifen | 75 | 14 | 0.62 (0.30-1.23) | 0.235 |
| | | No Tamoxifen | 83 | 22 | 1 (ref.) | |
| Neutrophils | Low | Tamoxifen | 80 | 7 | **0.19 (0.08-0.45)** | **0.001** |
| | | No Tamoxifen | 78 | 25 | 1 (ref.) | |
| | Inter. | Tamoxifen | 86 | 12 | **0.37 (0.18-0.76)** | **0.018** |
| | | No Tamoxifen | 72 | 21 | 1 (ref.) | |
| | High | Tamoxifen | 85 | 18 | 0.92 (0.45-1.90) | 0.841 |
| | | No Tamoxifen | 73 | 17 | 1 (ref.) | |
| T cells CD4 | Low | Tamoxifen | 85 | 9 | **0.21 (0.10-0.48)** | **0.001** |
| | | No Tamoxifen | 73 | 24 | 1 (ref.) | |
| | Inter. | Tamoxifen | 85 | 12 | 0.51 (0.25-1.06) | 0.122 |
| | | No Tamoxifen | 73 | 20 | 1 (ref.) | |
| | High | Tamoxifen | 81 | 16 | 0.74 (0.37-1.48) | 0.470 |
| | | No Tamoxifen | 77 | 19 | 1 (ref.) | |
| T cells CD8 | Low | Tamoxifen | 92 | 11 | **0.22 (0.10-0.47)** | **0.001** |
| | | No Tamoxifen | 66 | 22 | 1 (ref.) | |
| | Inter. | Tamoxifen | 81 | 13 | 0.52 (0.26-1.03) | 0.107 |
| | | No Tamoxifen | 77 | 23 | 1 (ref.) | |
| | High | Tamoxifen | 78 | 13 | 0.73 (0.34-1.55) | 0.485 |
| | | No Tamoxifen | 80 | 18 | 1 (ref.) | |
| T cells gamma delta | Low | Tamoxifen | 89 | 9 | **0.24 (0.11-0.54)** | **0.003** |
| | | No Tamoxifen | 69 | 21 | 1 (ref.) | |
| | Inter. | Tamoxifen | 90 | 14 | 0.44 (0.22-0.90) | 0.052 |
| | | No Tamoxifen | 68 | 20 | 1 (ref.) | |
| | High | Tamoxifen | 72 | 14 | 0.74 (0.37-1.46) | 0.472 |
| | | No Tamoxifen | 86 | 22 | 1 (ref.) | |
| T regulatory cells | Low | Tamoxifen | 87 | 9 | **0.27 (0.12-0.60)** | **0.005** |
| | | No Tamoxifen | 71 | 21 | 1 (ref.) | |
| | Inter. | Tamoxifen | 82 | 14 | 0.38 (0.19-0.76) | 0.018 |
| | | No Tamoxifen | 76 | 24 | 1 (ref.) | |
| | High | Tamoxifen | 82 | 14 | 0.79 (0.38-1.66) | 0.604 |
| | | No Tamoxifen | 76 | 18 | 1 (ref.) | |
| Macrophages M1 | Low | Tamoxifen | 88 | 6 | **0.13 (0.05-0.35)** | **0.001** |
| | | No Tamoxifen | 70 | 23 | 1 (ref.) | |
| | Inter. | Tamoxifen | 80 | 16 | 0.71 (0.37-1.37) | 0.398 |
| | | No Tamoxifen | 78 | 22 | 1 (ref.) | |
| | High | Tamoxifen | 83 | 15 | 0.66 (0.33-1.33) | 0.333 |
| | | No Tamoxifen | 75 | 18 | 1 (ref.) | |
| Macrophages M2 | Low | Tamoxifen | 85 | 9 | **0.20 (0.09-0.43)** | **0.001** |
| | | No Tamoxifen | 73 | 27 | 1 (ref.) | |
| | Inter. | Tamoxifen | 75 | 13 | 0.55 (0.27-1.09) | 0.139 |
| | | No Tamoxifen | 83 | 24 | 1 (ref.) | |
| | High | Tamoxifen | 91 | 15 | 0.93 (0.42-2.06) | 0.854 |
| | | No Tamoxifen | 67 | 12 | 1 (ref.) | |
| Monocytes | Low | Tamoxifen | 84 | 9 | **0.19 (0.08-0.45)** | **0.001** |
| | | No Tamoxifen | 74 | 24 | 1 (ref.) | |
| | Inter. | Tamoxifen | 87 | 9 | **0.28 (0.11-0.72)** | **0.020** |
| | | No Tamoxifen | 71 | 18 | 1 (ref.) | |
| | High | Tamoxifen | 80 | 19 | 0.90 (0.47-1.72) | 0.805 |
| | | No Tamoxifen | 78 | 21 | 1 (ref.) | |
| Plasma cells | Low | Tamoxifen | 90 | 14 | **0.23 (0.12-0.47)** | **0.001** |
| | | No Tamoxifen | 68 | 27 | 1 (ref.) | |
| | Inter. | Tamoxifen | 79 | 10 | 0.51 (0.23-1.17) | 0.164 |
| | | No Tamoxifen | 79 | 16 | 1 (ref.) | |
| | High | Tamoxifen | 82 | 13 | 0.48 (0.23-1.00) | 0.097 |
| | | No Tamoxifen | 76 | 20 | 1 (ref.) | |

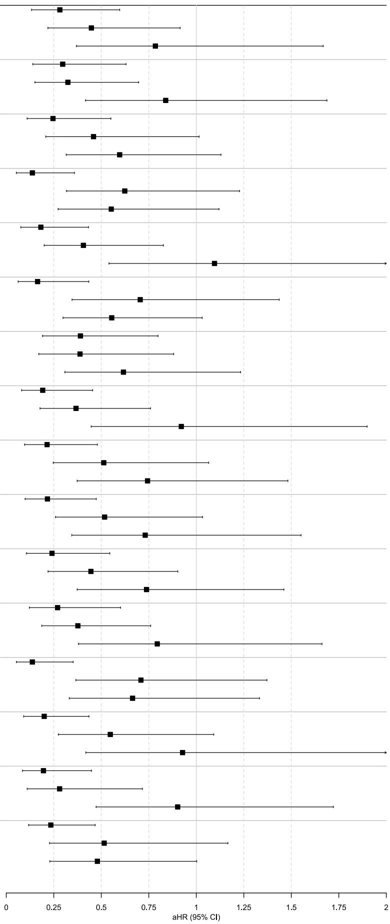


**Supplementary Figure S3:** Multivariable Cox proportional hazard regression analysis of DRFI in tamoxifen-treated patients compared to no tamoxifen treated patients across cell types in the TME. Multivariable analyses were adjusted for clinical information and tumour characteristics, including age, tumour size, tumour grade, progesterone receptor status, and Ki-67 status, and the administration of radiotherapy. DRFI, distant recurrence-free interval; DR, distant recurrence; aHR, adjusted hazard ratio; Ref, reference.
