## Supplemental Figure 4 for "The tumour microenvironment influences long-term tamoxifen benefit in postmenopausal ER+/HER2- breast cancer patients"

A

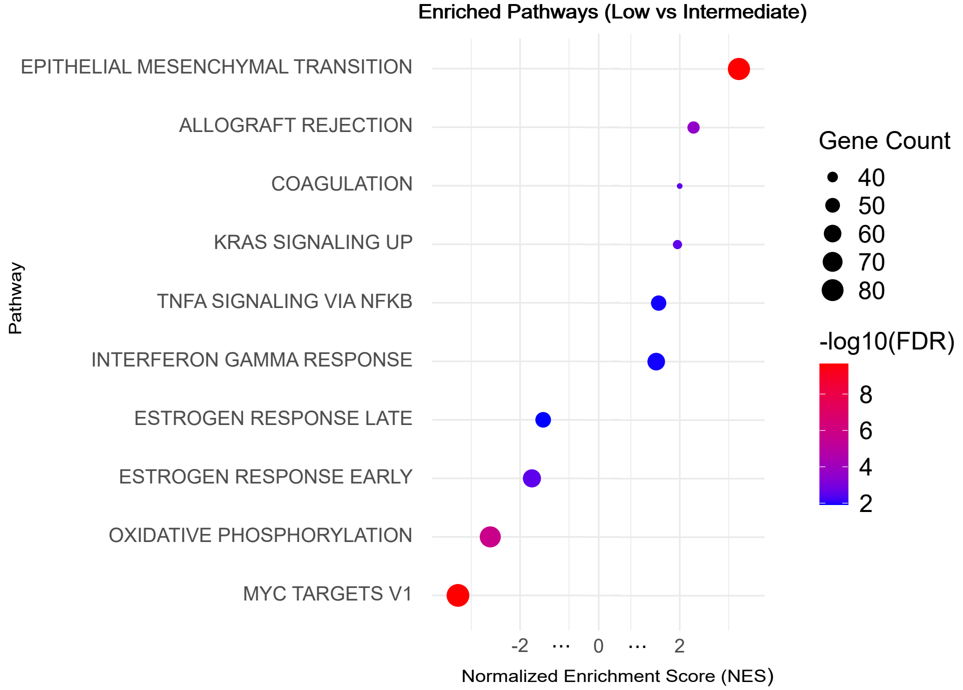


B

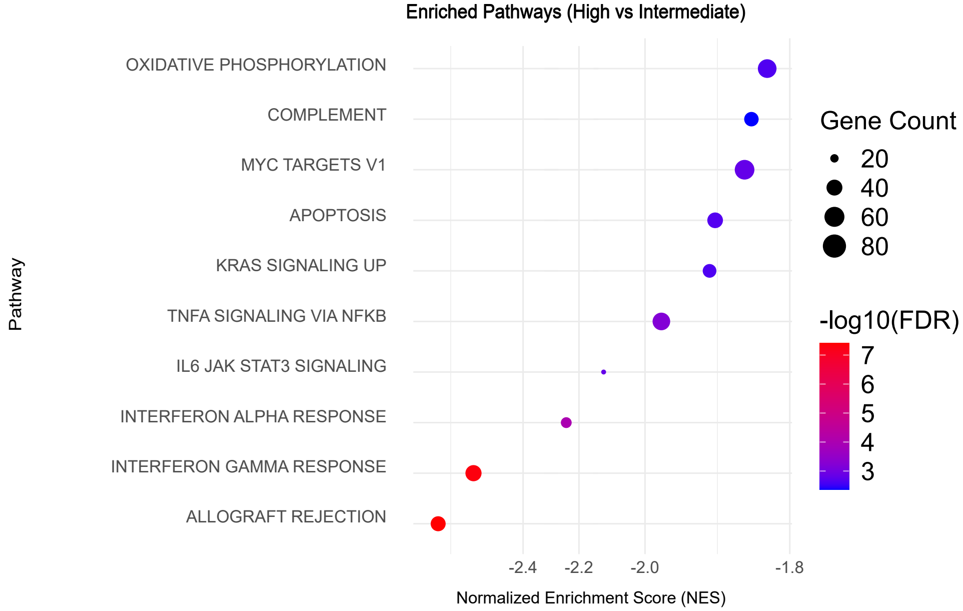


**Supplementary Figure S4**. Dot plot representation of the most and least enriched pathways for endothelial cells comparing (A) Low vs Intermediate scores, and (B) Intermediate vs High scores. The circle's size represents the number of genes in each pathway, and the colour of the qscore for each pathway.
